# Proteomic markers linking anxiety phenotypes to Alzheimer’s disease risk: a multi-analyte proteomic analysis

**DOI:** 10.64898/2026.09.16.26363134

**Authors:** Ana Paula Costa, Helmet T. Karim, Marissa F. Farinas, Mary M. Geissinger, Xuemei Zeng, Dana L. Tudorascu, Meryl A. Butters, Thomas K. Karikari, Carmen Andreescu

## Abstract

**Importance:** Anxiety phenotypes - rumination, global anxiety, and worry (RAW) - are increasingly associated with Alzheimer’s disease and related dementias (ADRD) risk, yet how anxiety phenotypes relate to coordinated, multi-system biology in the at-risk older adults is unknown.

**Objective:** To map associations between anxiety phenotypes and plasma proteomic profile across five ADRD-relevant biological domains in older adults with high anxiety symptoms.

**Design, Setting, and Participants:** Cross-sectional analysis of 110 older adults (aged 53-75 years) without dementia from the observational longitudinal RAW Brain study in Pittsburgh, Pennsylvania.

**Main Outcome(s) and Measure(s):** Plasma was profiled with the NULISAseq CNS panel. Principal component (PC) analysis was conducted with Horn’s parallel analyses across five ADRD-relevant biological domains (amyloid/tau pathology, inflammation, neurodegeneration, synuclein/synaptic disorders, and vascular) yielded 14 PCs (37.4-64.9% within-domain variance). Associations with each phenotype were tested by linear regression adjusted for age and sex, with Benjamini-Hochberg false discovery rate correction (p-FDR<0.05).

**Results:** A neurodegeneration component (higher neuronal pentraxins and neurofilament light, with lower neurogranin) was associated with all three phenotypes (rumination, β=0.26; global anxiety, β=0.39; worry, β=0.36; all p-FDR<0.05). Inflammatory, VEGF receptor-ligand, and phospho-tau versus amyloid-processing components showed phenotype-specific associations, most often with worry severity.

**Conclusion and Relevance:** Distinct anxiety phenotypes map onto dissociable plasma proteomic profiling, converging on a shared neurodegeneration axis reflecting neuronal injury, while diverging across neuroimmune, vascular, and proteostatic domains. These results point toward the biological heterogeneity of anxiety phenotypes as risk factors for ADRD. Further research is needed to clarify these biological pathways and explore targeted prevention strategies.

## INTRODUCTION

In older adults, anxiety is increasingly associated with cognitive decline, Alzheimer’s disease (AD), and other age-related dementias^1,2^. Clinically significant anxiety symptoms affect up to 15% of community-dwelling older adults and up to 28% of those in clinical settings^3,4^, and reported rates are higher still among individuals with subjective cognitive decline and mild cognitive impairment (MCI)^5^. Critically, because much of this evidence is derived from cross-sectional studies, it remains unclear whether anxiety constitutes a risk factor for cognitive decline^6,7^, an early manifestation of it^1,8,9^, or both. Beyond its association with dementia onset, anxiety has also been linked to more rapid disease progression^1,10^, suggesting it may be relevant across the full trajectory of cognitive aging.

Although several studies link anxiety to an increased risk of dementia, independently of depression^11,12^, with meta-analyses estimating 77% greater risk of cognitive impairment^1^, 53% risk of Alzheimer’s disease^13^, and 65% risk of vascular dementia^14^, findings across studies have been inconsistent, with differences observed by cognitive stage, dementia subtype, and study population. This clinical heterogeneity has motivated efforts to characterize the association at the biomarker level. In older adults, we have shown that worry and rumination severity track with age-related neurobiological changes, including cortical thickness reductions^15^, accelerated brain aging^16^, lower hippocampal subfield volume^17^, and altered connectivity in stress-and emotion-processing circuits^18,19^. Molecular imaging extends these findings to AD proteinopathy. Among more than 4,400 participants in the A4/LEARN studies, greater amyloid-β (Aβ) deposition - but not tau - was associated specifically with worry rather than with global anxiety^20^. Emerging high-sensitivity blood assays now permit simultaneous quantification of neurodegenerative, inflammatory, and Alzheimer’s disease and related dementia (ADRD) biomarkers, including Aβ, phosphorylated tau (p-tau), neurofilament light (NfL), and glial fibrillary acidic protein (GFAP) in relation to neuropsychiatric symptoms. Yet these studies have examined single analytes in isolation^5^. It remains unresolved how anxiety phenotypes relate to the coordinated, multi-system biology of the preclinical period, and whether such associations are shared across phenotypes.

Multi-analyte plasma proteomics profiling offers a tractable way to address this, permitting simultaneous, minimally invasive quantification of hundreds of ADRD-relevant proteins spanning amyloid and tau pathologies, neurodegeneration, inflammation, vascular, and synuclein/synaptic disorders in cognitively unimpaired older adults^21^. This approach makes it possible to characterize not only domain-specific associations but also the inter-domain relationships that emerge across biological systems, providing a more integrated view than imaging or single-analyte studies allow.

In the present study, we used a multi-analyte plasma proteomic analysis in the RAW Brain cohort to examine how anxiety phenotypes - rumination, global anxiety, and worry (RAW) - relate to ADRD-relevant biology across five domains in older adults. We had three aims: (1) to characterize the cross-domain structure of the proteomic data by constructing an inter-domain correlation network to identify interpretable biological axes; (2) to test associations between each biological domain and each anxiety phenotype; and (3) to examine whether these associations were shared across anxiety phenotypes or are specific to individual ones. Given the cross-sectional analysis, we framed all relationships as associations rather than as evidence of causal or mechanistic pathways, and we additionally explore whether associations varied by time from symptom onset.

## METHODS AND MATERIALS

### Participants and study design

The study cohort consisted of participants from the RAW Brain study (R01 MH108509 The RAW Brain – The Effect of Rumination, Anxiety and Worry on Aging and Dementia Risk) in Pittsburgh, Pennsylvania, USA. The RAW Brain study is an ongoing observational longitudinal study with visits 2 years apart including demographic, clinical, behavioral, plasma biomarkers, and neuroimaging data collection; aimed at understanding of the relationships between late-life anxiety and ADRD risk. Participants >50 years of age were recruited through a web-based recruitment resource at the University of Pittsburgh (Pitt+Me), in-person referrals, flyers, and radio/television ads. For this study, we included the baseline assessment of 110 participants. Participants were recruited using key dimensions of anxiety - rumination, global anxiety, worry (RAW) - this dimensional design allowed us to tap into an understudied yet highly prevalent population of older participants with severe rumination, anxiety or worry, that may or may not qualify for a categorical diagnosis^22,23^. Participants could have a diagnosis of anxiety disorder (e.g., generalized anxiety disorder, panic disorder, etc.) and/or depressive disorders (e.g., major depressive disorder, persistent depressive disorder, etc.), but that was not required. Exclusion criteria for the study were 1) other major psychiatric disorder including any form of psychosis, bipolar disorder, autism spectrum disorder, and intellectual development disorder; 2) dementia [Modified Mini-Mental State Examination score <84]; 3) high suicide risk; 4) use of selective serotonin reuptake inhibitor or serotonin and norepinephrine reuptake inhibitor antidepressants that participants were not willing/able to discontinue before the scan (5-to 14-day washout depending on the antidepressant, 6 weeks for fluoxetine); 5) history of drug or alcohol abuse within the last 6 months; 6) high doses of benzodiazepines (≥2 mg of lorazepam/day); 7) uncorrected vision problems that preclude testing; 8) below sixth grade reading level; 9) clinical diagnosis of cerebrovascular accident, multiple sclerosis, vasculitis, or significant head trauma; and 10) contraindications to MRI. The University of Pittsburgh Institutional Review board approved the study. All participants provided written informed consent.

### Anxiety phenotype classification and measures

Along with demographic information (age, sex, race, ethnicity, and education), we assessed the following: worry severity (PSWQ, Penn State Worry Questionnaire^24,25^), global anxiety (HARS, Hamilton Anxiety Rating Scale^26^), and rumination (RSQ, Rumination Subscale from the Response Style Questionnaire^27^). Participants were classified as High-RAW (n=48) if they meet thresholds on ≥1 domain: PSWQ ≥50 and/or RSQ >50, and/or HARS >17. Participants not meeting any criterion were classified as Low-RAW (n=62). For the primary regression analyses, all three measures were treated as continuous variables to preserve statistical power and capture dimensional variation in anxiety phenotypes.

Participants were further characterized clinically using measures of depression (HAMD, Hamilton Depression Rating Scale), neuroticism (FFI, Five-Factor Inventory), perceived stress (Perceived Stress Scale – 10 item), medical burden (CIRS-G, Cumulative Illness Rating Scale – Geriatrics), functional impairment (WHODAS 2.0, World Health Organization Disability Assessment Schedule II-36 Items), lifetime history of psychiatric disorders as per DMS-5 criteria (MINI, Mini International Neuropsychiatric Interview) as well as a comprehensive neuropsychological assessment. Global cognitive function was assessed with the Repeatable Battery for the Assessment of Neuropsychological Status (RBANS) and the Montreal Cognitive Assessment (MoCA). The RBANS is a standardized, age-normed test that assesses five domains including immediate and delayed memory, visuospatial ability, language function, and attention and yields a Total Index Score. The MoCA is a 30-point cognitive screener to detect dementia for clinical care and research, it consists of 12 brief tasks covering visuospatial/executive function, naming, memory, attention, language, delayed recall, and orientation that yield a single total score.

### Blood collection and processing

Blood samples were collected according to the procedures described in Zeng et al^28^. Briefly, blood was collected via venipuncture by experienced nurses trained at Western Psychiatric Hospital-UPMC Clinic. Whole blood from each participant was collected into a 10 mL lavender top EDTA tube. Following each blood draw, the EDTA tubes were promptly inverted 8-10 times and centrifuged at 2,000 *x g* for 10 minutes at 4°C. The resulting plasma samples were aliquoted into cryovials and frozen at −80°C until use.

### NULISAseq assay procedures and data processing

Plasma samples were thawed and centrifuged at 10,000 *x g* for 10 min to remove particulates. The supernatants were analyzed using the NUcleic acid Linked Immuno-Sandwich Assay (NULISA), a recently developed ultrasensitive multiplex immunoassay that enables the simultaneous detection of many low-abundance proteins^29^. Samples were analyzed using one of its multiplex panels, the NULISAseq™ CNS Disease Panel 120 (hereafter referred to as the NULISAseq CNS panel) on an Alamar ARGO HT system as previously described^21^. Briefly, samples were incubated with a cocktail of paired capture and detection antibodies and internal control mCherry protein. The capture antibodies are conjugated with partially double-stranded DNA containing a poly-A tail and a target-specific barcode while the detection antibodies are conjugated with another partially double-stranded DNA containing a biotin group and a matching target-specific barcode. The immunocomplexes underwent magnetic bead-based capture, followed by washing, release into a low-salt buffer, recapture with streptavidin-coated magnetic beads, and a second round of washing. DNA reporter molecules containing unique target and sample-specific barcodes were then generated by ligation and quantified using next generation sequencing (NGS). A total of 131 biomarkers were included in the NULISAseq™ CNS Disease Panel 120 pre-classified into a prior five biologically Alzheimer’s disease and related dementias (ADRD) domains: Amyloid and Tau Pathologies, Inflammation, Neurodegeneration, Synuclein and Synaptic Disorders, and Vascular (**Supplementary Table S1**). This domain structure was encoded in a companion mapping file that linked each protein target name to its biological category and was used to organize all downstream dimension-reduction analyses. Protein levels for each target were quantified by first normalizing the raw counts. This was achieved by dividing the target count for each sample well (plus one) by the corresponding mCherry internal control count (plus one). The resulting values were then rescaled and log₂-transformed to generate NULISA Protein Quantification (NPQ) units, which served as surrogate measures of protein abundance. Fold changes were calculated as 2 to the power of the difference in NPQ. A sample control was measured in triplicate to assess the reproducibility of the assay.

### Statistical analysis

To characterize the panel’s global co-expression architecture, we computed pairwise Pearson correlations between all proteins. Principal component analysis (PCA) was performed separately within each of the five biological domains to reduce dimensionality while preserving domain-specific structure. Components retained per domain were determined by Horn’s parallel analysis (5,000 Monte Carlo interactions, α=0.05, seed=123). To assess variation shared across domains, we computed Pearson correlations between all pairs of PC scores across the 14 retained components and applied Benjamini-Hochberg (BH) false discovery rate (FDR) correction. Correlations with FDR-adjusted p≤0.0021 were considered significant and shown in the cross-domain network (**Supplementary Table S4 and Figure S1**).

Multiple linear regression model assumptions were examined via residuals and influential points. No departures from normality were observed or outliers. Models were fitted for each pairing of PC score (14 PCs) and anxiety phenotype (worry, global anxiety, rumination) adjusting for age and sex, yielding 42 models (14 PCs x 3 phenotypes. All continuous variables (PC scores and outcome) were z-standardized, enabling interpretation of regression coefficients as standardized effect sizes (standardized β). Each model reported the β and p-value. BH-FDR correction was applied within each outcome across all 14 PC tests (FDR-corrected p< 0.05).

To test whether associations varied by symptom chronicity, all regressions were repeated within two subgroups defined by self-reported duration of symptom onset (<10 vs ≥10 years). Covariate adjustment (age and sex), and standardization matched the primary analysis, and BH-FDR correction was applied within each group and outcome. Given the reduced subgroup sample sizes, these results are interpreted as exploratory and hypothesis-generating. Analyses were performed in R, using the *dplyr, gtsummary, broom, bestNormalize, effectsize*, and *pwr* packages.

## RESULTS

### Participant characteristics

Demographics and clinical characteristics of participants are shown in **Table 1**. A total of 110 participants from The RAW Brain study were included in the multi-analyte plasma proteomic profiling. The High-RAW group (n= 48) was younger than the Low-RAW group (n= 62). Groups did not differ significantly in sex, race, ethnicity, years of education, or APOEε4 genotype. By design, the High-RAW group evidenced markedly elevated scores on all three anxiety phenotype measures compared to the Low-RAW group on PSWQ, RSQ, and HARS. The RBANS total score and MoCA total score were not significantly different between High-and Low-RAW groups.

**Table 1.** Demographics and Clinical Characteristics of Participants.

| **Table 1.** Demographics and Clinical Characteristics of Participants |  |  |  |  |
| --- | --- | --- | --- | --- |
| **Variable** | **Low-RAW**<br>**N = 62<sup>1</sup>** | **High-RAW**<br>**N = 48<sup>1</sup>** | **p-value<sup>2</sup>** | **Overall**<br>**N = 110<sup>1</sup>** |
| **Age, years, mean (SD)** | 67.1 ± 8.5 | 61.6 ± 7.8 | **0.001** | 64.7 ± 8.6 |
| **Sex, n, %** |  |  | 0.400 |  |
| Male | 24 (39%) | 14 (29%) |  | 38 (35%) |
| Female | 38 (61%) | 34 (71%) |  | 72 (65%) |
| **Race, n, %** |  |  | >0.999 |  |
| White | 53 (85%) | 41 (85%) |  | 94 (85%) |
| Non-White | 9 (15%) | 7 (15%) |  | 16 (15%) |
| **Ethnicity, n, %** |  |  | 0.592 |  |
| Not Hispanic or Latino | 60 (97%) | 48 (100%) |  | 108 (98%) |
| Hispanic or Latino | 2 (3.2%) | 0 (0%) |  | 2 (1.8%) |
| **APOE4 carrier status, n, %** |  |  | 0.568 |  |
| Non-carrier | 41 (76%) | 38 (83%) |  | 79 (79%) |
| Carrier | 13 (24%) | 8 (17%) |  | 21 (21%) |
| **Education, years, mean (SD)** | 16.8 ± 2.2 | 16.3 ± 2.2 | 0.329 | 16.6 ± 2.2 |
| **MoCA total, mean (SD)** | 27.1 ± 2.1 | 27.0 ± 2.3 | 0.897 | 27.1 ± 2.2 |
| **RBANS total, mean (SD)** | 107.8 ± 11.5 | 108.3 ± 14.1 | 0.757 | 108.0 ± 12.6 |
| **PSWQ (Worry), mean (SD)** | 32.2 ± 9.0 | 57.9 ± 11.2 | **<0.001** | 43.4 ± 16.2 |
| **RSQ (Rumination), mean (SD)** | 29.6 ± 5.6 | 48.2 ± 11.8 | **<0.001** | 37.7 ± 12.8 |
| **HARS (Anxiety), mean (SD)** | 5.5 ± 3.9 | 15.1 ± 7.1 | **<0.001** | 9.7 ± 7.3 |

<sup>1</sup> Mean ± SD; n (%)
<sup>2</sup> Wilcoxon rank sum test; Pearson's Chi-squared test
Mean ± SD for continuous variables; n (%) for categorical variables.
p-values: Wilcoxon rank-sum test (continuous); Pearson Chi-squared test (categorical).

### Cross-domain principal components correlation network

**Figure 1B** displays cross-correlations among proteins in the NULISAseq CNS panel. Plasma biomarkers were organized into five a priori biological domains and subjected to principal component analysis (PCA) within each domain, with the number of retained components determined by Horn’s parallel analysis. Across domains, PCA yielded 14 principal components (PCs), collectively explaining 37.4-64.9% of within-domain variance (**Figure 1C**; **Supplementary Table S2**). Component loadings for all domains are presented in **Supplementary Table S3**. The resulting correlation network revealed substantial inter-domain coupling across the five biological domains (**Figure 2**), particularly among neurodegeneration, neuroinflammation, and vascular biomarkers. The network identified five interpretable biological axes: (1) phospho-tau versus amyloid-processing axis; (2) lymphocyte priming versus inflammation axis; (3) synaptic plasticity versus neuronal activity marker axis; (4) protein aggregation and neuronal loss axis; and (5) vascular integrity versus ligand-driven angiogenic axis (**Supplementary Figure S2A-E; Supplementary results**). These inter-domain correlations motivated the domain-stratified PCA approach by demonstrating that while domains share variance, they retain distinct biological signals warranting separate characterization.

**Figure 1.**
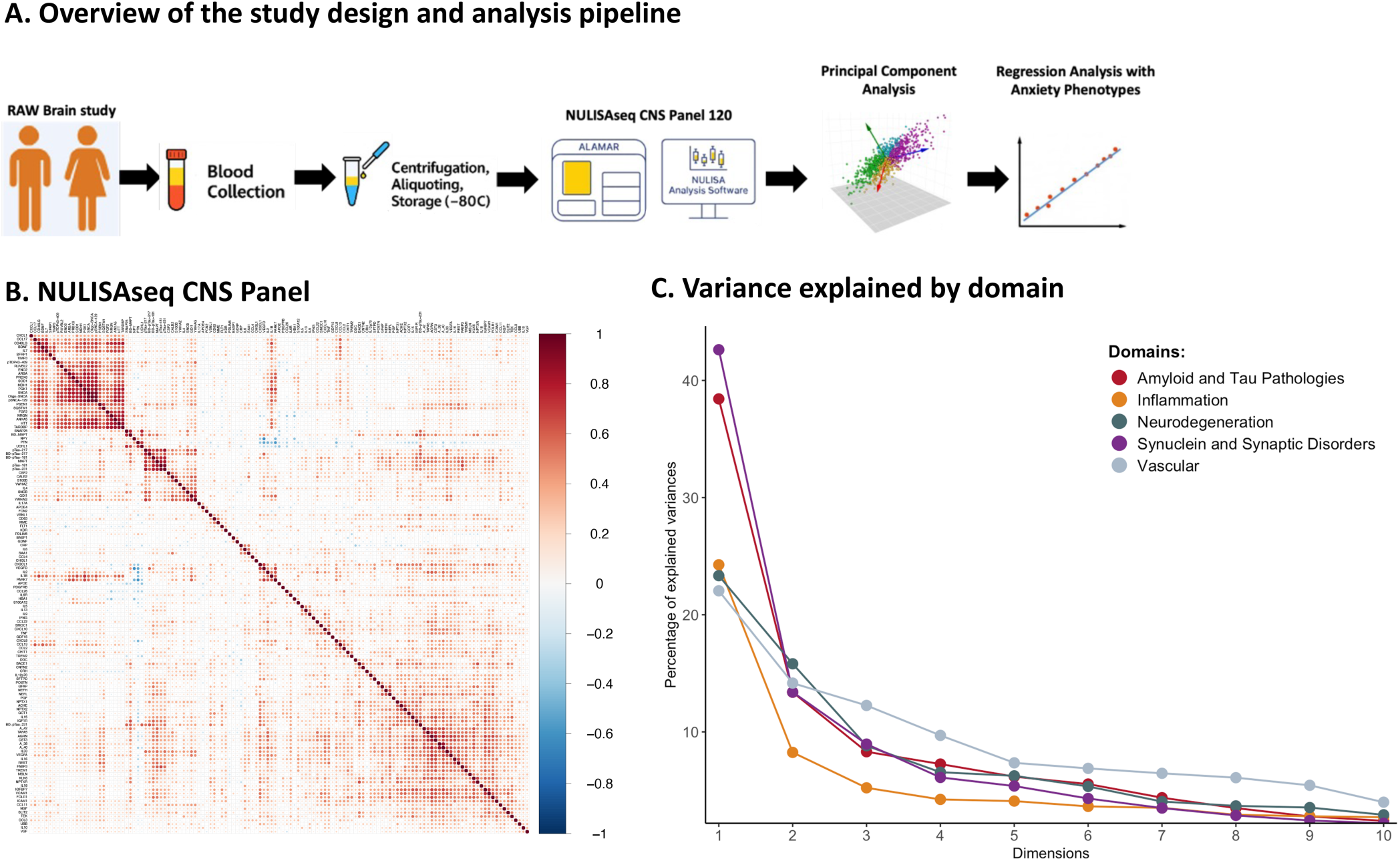
Principal component analysis of NULISAseq CNS panel in the cross-sectional The RAW Brain cohort. **A)** Overview of the study design and analysis pipeline. **B)** Correlation heatmap for NULISAseq CNS panel plasma proteins. **C)** Screen plot displaying variance explained by each principal component from Principal Component Analysis (PCA) by domain.

**Figure 2.**
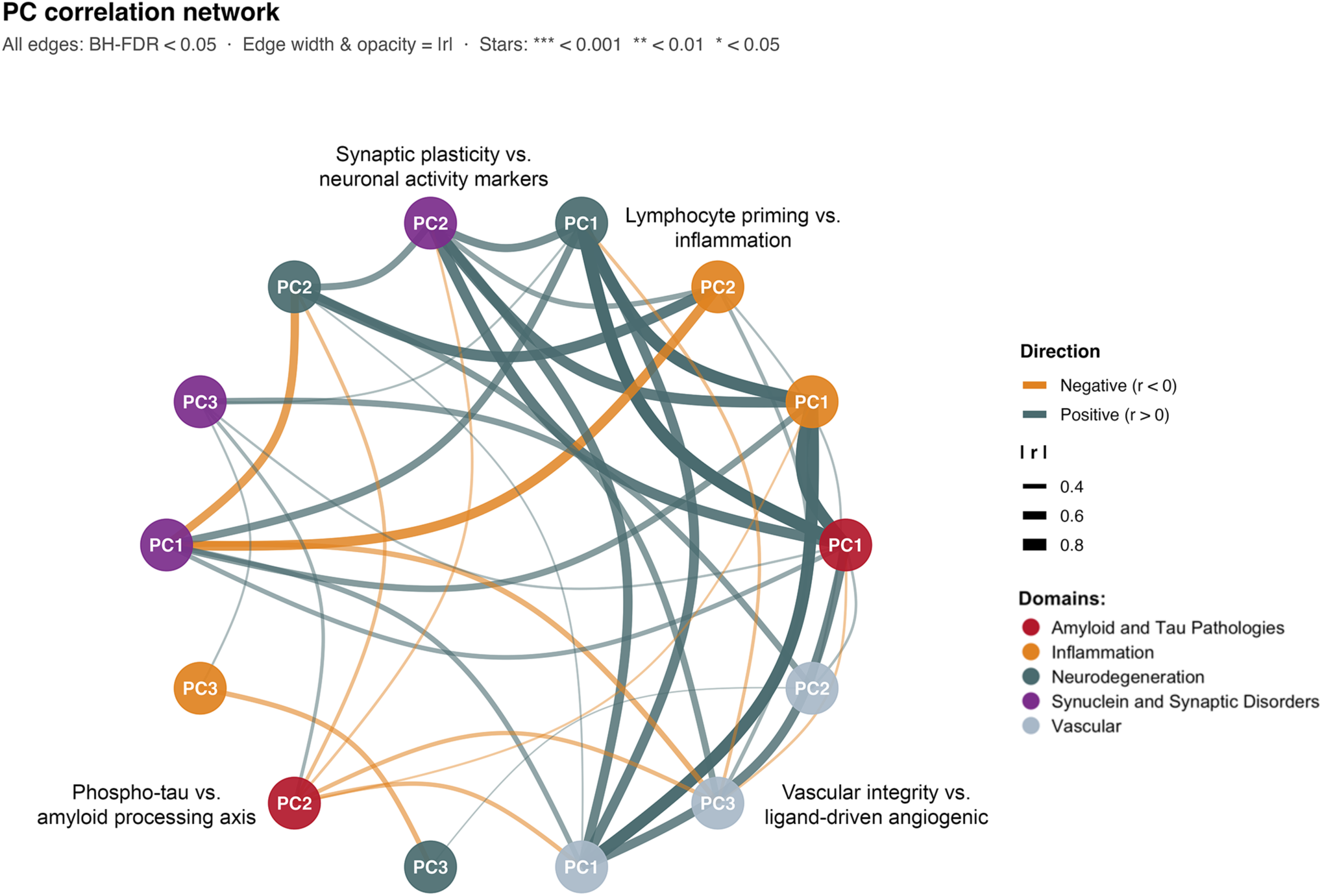
Cross-domain principal component correlation network of plasma proteomics in a cohort of older adults enriched for anxiety phenotypes. Nodes represent the 14 principal components (PCs), colored by domain. Edges are pairwise Pearson correlations significant after Benjamini-Hochberg correction (p-FDR≤0.0021). Edge color denotes direction (teal, positive; orange, negative) and edge width/opacity scale with |r|. FDR-adjusted significance (*p<0.05, **p<0.01, ***p<0.001). PCs associated with anxiety phenotypes: Neurodegeneration PC2: Synaptic plasticity vs. neuronal activity markers, Inflammation PC1: Lymphocyte priming vs. inflammation, Amyloid and tau pathologies PC2: Phospho-tau vs. amyloid processing axis, Vascular PC3: Vascular integrity vs. ligand-driven angiogenic.

We provide an integrated overview of these associations with a focus on those associated with anxiety phenotypes. Cross-correlations analysis revealed a densely interconnected network, with 9 of the 10 possible pairwise associations significant after FDR correction; only the phospho-tau vs. amyloid-processing axis (Amyloid and Tau Pathologies PC2) and the protein aggregation and neuronal loss axis (Synuclein and Synaptic Disorders PC1) were not significantly correlated. Notable, the synaptic plasticity vs. neuronal activity marker axis (Neurodegeneration PC2) emerged as a central hub, correlating with all four other axes: greater synaptic disorganization was correlated with a more inflammatory peripheral profile (Inflammation PC2; r=-0.74, q<0.001), greater protein aggregation and neuronal-loss burden (r=0.60, q<0.001), greater VEGF receptor-ligand imbalance (Vascular PC3; r=0.51, q<0.001), and lower phospho-tau relative to amyloid-processing markers (r=0.34, q<0.01). These relationship extended across the broader network: a more inflammatory profile also corresponded to greater aggregation and neuronal-loss burden (r=-0.69, q<0.001), greater VEGF receptor-ligand imbalance (r=-0.35, q<0.001), and lower phospho-tau relative to amyloid-processing markers (r=-0.26, q<0.05), and greater VEGF receptor-ligand imbalance corresponded to both greater aggregation burden (r=0.44, q<0.001) and lower phospho-tau relative to amyloid-processing markers (r=0.37, q<0.001).

### Principal component associations with anxiety phenotypes

Across the three anxiety phenotypes (rumination, global anxiety, and worry; RAW) and 14 PCs (42 tests), FDR-corrected analyses identified 10 statistically significant associations (p-FDR<0.05; **Figure 3A-D**, **Supplementary Figure S3 and Table 2**). Results of the associations are summarized in **Supplementary Tables S5-S7**.

**Figure 3.**
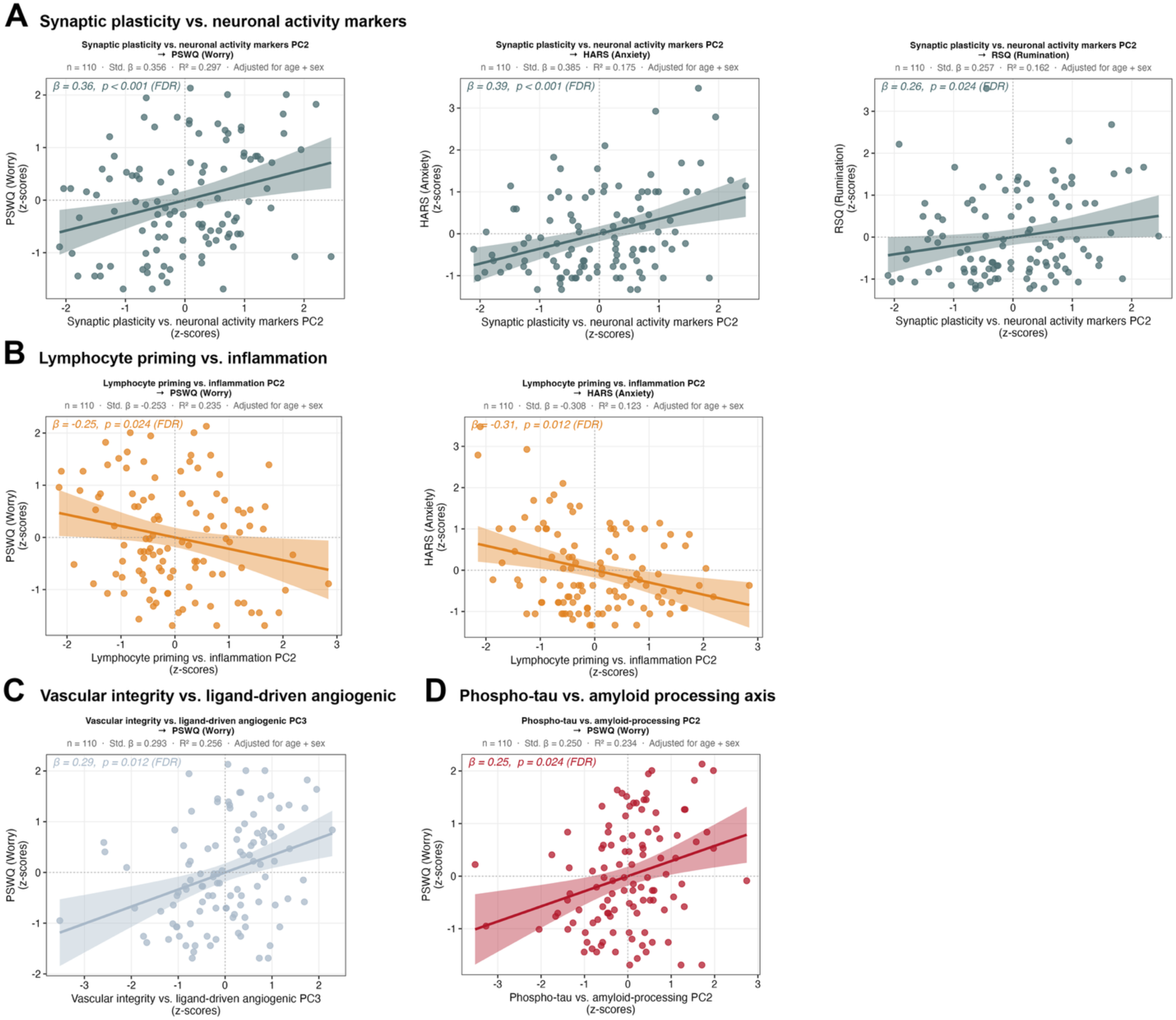
Anxiety-associated principal components (PCs) and their cross-domain correlates. Each panel (A–D) corresponds to one biological axis significantly associated with one or more anxiety phenotypes. Scatterplots of the PCs against anxiety phenotype, with linear regression fit (solid line) and 95% confidence band (shaded). All models were adjusted for age and sex (N = 110); standardized β, FDR-adjusted p-value, and model R^2^ are shown.

**Table 2.**
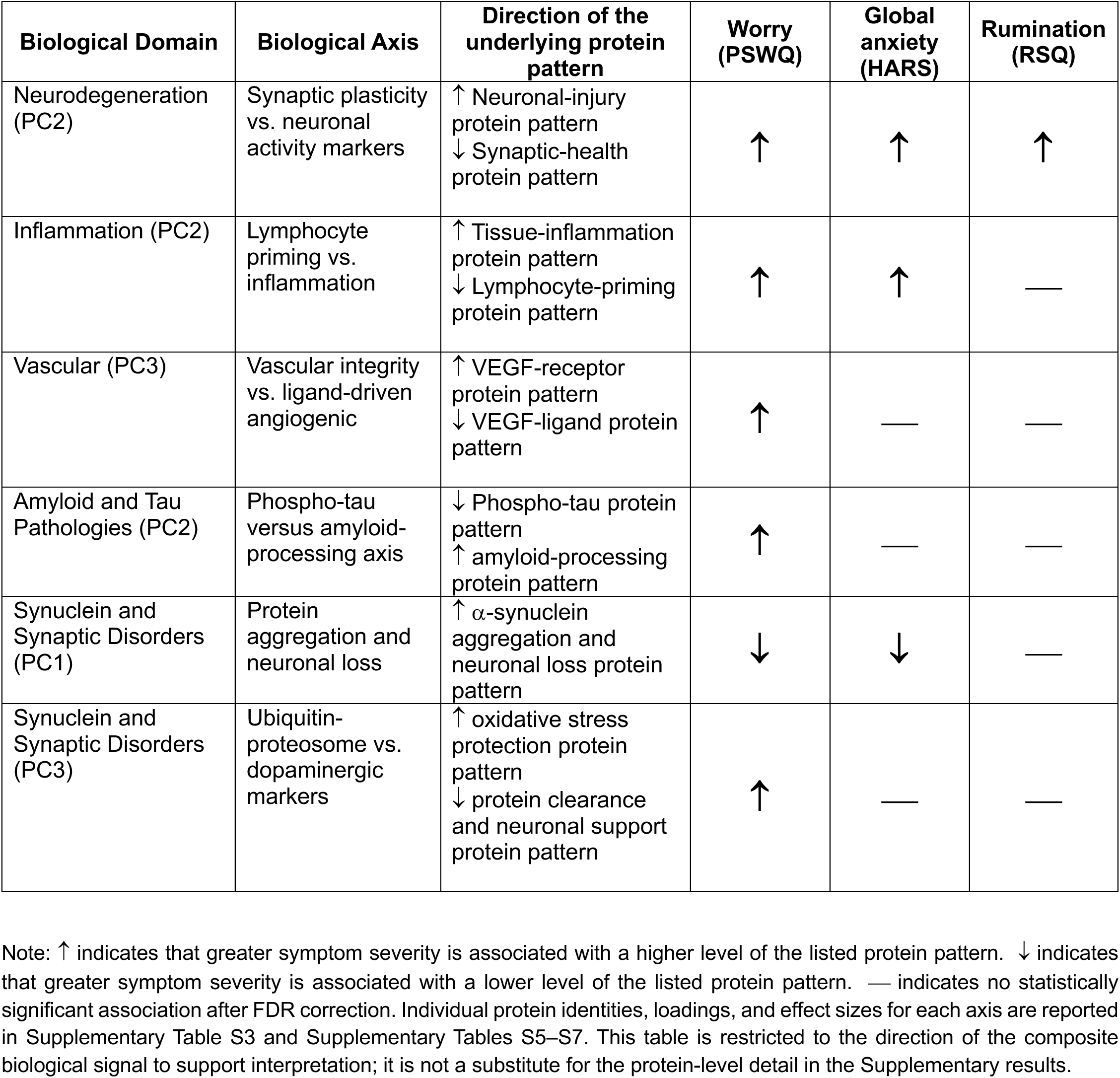
Associations between anxiety phenotypes and ADRD-relevant proteomic domains.

| Biological Domain | Biological Axis | Direction of the underlying protein pattern | Worry (PSWQ) | Global anxiety (HARS) | Rumination (RSQ) |
| --- | --- | --- | --- | --- | --- |
| Neurodegeneration (PC2) | Synaptic plasticity vs. neuronal activity markers | ↑ Neuronal-injury protein pattern<br>↓ Synaptic-health protein pattern | ↑ | ↑ | ↑ |
| Inflammation (PC2) | Lymphocyte priming vs. inflammation | ↑ Tissue-inflammation protein pattern<br>↓ Lymphocyte-priming protein pattern | ↑ | ↑ | — |
| Vascular (PC3) | Vascular integrity vs. ligand-driven angiogenic | ↑ VEGF-receptor protein pattern<br>↓ VEGF-ligand protein pattern | ↑ | — | — |
| Amyloid and Tau Pathologies (PC2) | Phospho-tau versus amyloid-processing axis | ↓ Phospho-tau protein pattern<br>↑ amyloid-processing protein pattern | ↑ | — | — |
| Synuclein and Synaptic Disorders (PC1) | Protein aggregation and neuronal loss | ↑ α-synuclein aggregation and neuronal loss protein pattern | ↓ | ↓ | — |
| Synuclein and Synaptic Disorders (PC3) | Ubiquitin-proteasome vs. dopaminergic markers | ↑ oxidative stress protection protein pattern<br>↓ protein clearance and neuronal support protein pattern | ↑ | — | — |

Note: ↑ indicates that greater symptom severity is associated with a higher level of the listed protein pattern. ↓ indicates that greater symptom severity is associated with a lower level of the listed protein pattern. — indicates no statistically significant association after FDR correction. Individual protein identities, loadings, and effect sizes for each axis are reported in Supplementary Table S3 and Supplementary Tables S5–S7. This table is restricted to the direction of the composite biological signal to support interpretation; it is not a substitute for the protein-level detail in the Supplementary results.

#### Synaptic plasticity vs. neuronal activity markers is the most robust cross-dimensional correlate of anxiety phenotypes

Synaptic plasticity vs. neuronal activity marker axis (Neurodegeneration PC2) demonstrated the strongest and most consistent associations across all three anxiety phenotypes (**Figure 3A**). Higher scores were significantly associated with greater worry (Std.β=0.36; 95% CI, 0.19-0.52; p-FDR<0.001), higher global anxiety (Std.β=0.39; 95% CI, 0.21-0.56; p-FDR<0.001), and greater rumination (Std.β=0.26; 95% CI, 0.08-0.43; p-FDR=0.024).

Higher scores indexed higher plasma neuronal pentraxins (NPTX1, NPTX2, NPTXR), neurofilament light (NEFL), and visinin-like protein 1 (VSNL1), with lower neurogranin, annexin A5, FGF2, and enolase 2 (Supplementary results). Several of these, including NPTX1, NEFL, and VSNL1, have been reported as markers of synaptic and neuronal injury. Synaptic plasticity vs. neuronal activity marker axis was moderately correlated with the first two PCs of the Synuclein and Synaptic Disorders (PC1; r=0.60; PC2; r=-0.52; both p-FDR<0.001). Given the PCs loadings (Supplementary results), higher axis scores corresponded to lower levels of aggregation-prone α-synuclein and TDP-43 species and to higher levels of neurotrophic and synaptic markers with lower oligomeric α-synuclein. The axis was also weakly and negatively correlated with Vascular (PC1; r=-0.24; FDR<0.001), a direction corresponding to higher levels of VEGF-pathway, hypoxia-related, and inflammatory markers (**Supplementary Figure S2A**).

#### Lymphocyte priming versus inflammation axis shows inverse associations with worry and global anxiety

Lymphocyte priming versus tissue inflammation axis (Inflammation PC2) was inversely associated with worry (Std.β=−0.25; 95% CI, −0.42 to −0.08; p-FDR=0.024) and global anxiety (Std.β=−0.31; 95% CI, −0.49 to −0.13; p-FDR=0.012) (**Figure 3B**). Because higher scores on this axis index the lymphocyte-priming pattern and lower scores index the inflammatory pattern (Supplementary results), these inverse associations indicate that higher worry and anxiety corresponded to lower axis scores, that is, the peripheral inflammatory markers, with relatively higher VCAM, CX3CL1, CRH, IFN-γ, and GFAP and relatively lower CD40L, IL-7, CCL17, and IL-1β. This axis was also weakly and positively correlated with Vascular PC2 (r=0.26; p-FDR=0.01) (**Supplementary Figure S2B**), a direction corresponding to lower angiogenic and matrix-remodeling markers together with higher VEGFD and hypoxia-related markers.

#### Vascular integrity versus ligand-driven angiogenic axis associates selectively with worry

Vascular integrity vs. ligand-driven angiogenic axis (Vascular PC3) was significantly associated with worry (Std.β=0.29; 95% CI, 0.13,0.46; p-FDR=0.012) but not with rumination or global anxiety (**Figure 3C**). Higher scores on this axis index elevated VEGF receptors (FLT1 and KDR) with reduced VEGF ligands (VEGFA and PGF) and acute-phase inflammatory (SAA1) and matrix remodeling (POSTN) markers (Supplementary results). Higher worry therefore corresponded to this receptor-ligand imbalance in VEGF signaling. This axis was also positively correlated with the synaptic plasticity vs. neuronal activity marker axis (Neurodegeneration PC2; r=0.51; p-FDR<0.001) (**Supplementary Figure S2C**), indicating that the worry-associated vascular profile co-occurred with peripheral markers of early synaptic disorganization.

#### Phospho-tau versus amyloid-processing axis is associated with worry

Phospho-tau versus amyloid-processing axis (Amyloid and Tau Pathologies PC2) was associated with worry (Std.β=0.25; 95% CI, 0.08,0.42; p-FDR=0.024) but did not survive FDR correction for rumination or global anxiety, although it trended in the same direction (**Figure 3D**). Higher worry was associated with a biomarker profile of lower phospho-tau (p-tau181, p-tau217, and p-tau231) and higher Aβ40, Aβ38, and amyloid-processing markers (APOE, KLK6, and BACE1) (Supplementary results). The axis correlated with several domains, including positively with Vascular PC3 (r=0.37; p-FDR<0.001) and Neurodegeneration PC2 (r=0.34; p-FDR<0.01) (**Supplementary Figure S2D**). The positive correlations suggesting that this axis co-occurs with early synaptic disorganization alongside the VEGF receptor-ligand imbalance.

#### Protein aggregation and neuronal loss axis contribute to anxiety and worry

Protein aggregation and neuronal loss axis (Synuclein and Synaptic Disorders PC1), with higher scores reflecting greater burden (Supplementary results), was inversely associated with anxiety (Std.β=-0.27; 95% CI, −0.46 to −0.09; p-FDR=0.02) and worry (Std.β=-0.23; 95% CI, −0.40 to −0.06; p-FDR=0.04), and was not associated with rumination. Higher anxiety and worry therefore corresponded to lower levels of α-synuclein aggregation markers (phospho-Ser129, total, and oligomeric α-synuclein) and neuronal-loss markers. This component was correlated with two vascular components, Vascular PC1 (r=-0.43; p-FDR<0.001) and Vascular PC3 (r=-0.44; p-FDR<0.001) (**Supplementary Figure S2E**). Because Vascular PC1 is oriented with higher scores at the lower VEGF and hypoxia, the negative correlation indicates that greater aggregation burden co-occurred with higher VEGF-pathway and hypoxia-related markers. PC3 of the same domain (ubiquitin-proteasome and dopaminergic axis) was associated with worry (Std.β=0.26; 95% CI, 0.09-0.44; p-FDR=0.02) and was not associated with anxiety or rumination. Higher scores index higher PARK7/DJ-1 with lower UCHL1, β-synuclein, and NGF (Supplementary results), so higher worry corresponded to this PARK7-high, UCHL1-low pattern.

### Sensitivity and Supplementary Analyses

To investigate whether the primary plasma proteomic profiling associations differed by onset of symptoms, we stratified the participants by symptom onset <10 or ≥10 years ago. Three key divergences from the primary results are highlighted (**Supplementary Figure S4A and B**). First, the Synaptic plasticity vs. neuronal activity markers domain - the most robust primary finding across all three anxiety phenotypes - was selectively attenuated in both groups, most strikingly in the <10-year subgroup, suggesting that this synaptic disorganization axis may require a larger or more heterogeneous sample to be reliably detected within chronicity subgroups. Second, the direction of association between phospho-tau versus amyloid-processing axis and rumination reversed across groups, among individuals with recent symptom onset, higher rumination was associated with greater tau phosphorylation relative to amyloid, whereas the primary analysis showed the opposite.

## DISCUSSION

In this cross-sectional analysis of older adults at risk for Alzheimer’s disease (AD) and related dementias (ADRD), we integrated plasma proteomic profiling with measures of anxiety phenotypes to map their biological correlates across five ADRD-relevant domains. The most consistent association was between heightened RAW and the neurodegeneration-domain component (Neurodegeneration PC2, characterized by higher neuronal pentraxins and neurofilament light together with lower neurogranin). Of the 14 components examined, it was the only one associated with all three anxiety phenotypes after FDR correction. Beyond this component, three further axes showed selective associations, most often with more severe worry: a lymphocyte priming vs. inflammation axis (Inflammation PC2), a VEGF receptor-ligand axis (Vascular PC3), and a phospho-tau versus amyloid-processing axis (Amyloid and Tau Pathologies PC2). Collectively, these associations are consistent with a multi-pathway biological model of anxiety in the context of ADRD risk, in which neuronal injury profiling is the most consistent correlate across anxiety phenotypes while vascular, inflammatory, and amyloid-tau processes show dimension-specific associations (see **Table 2** and **Figure 4**).

**Figure 4.**
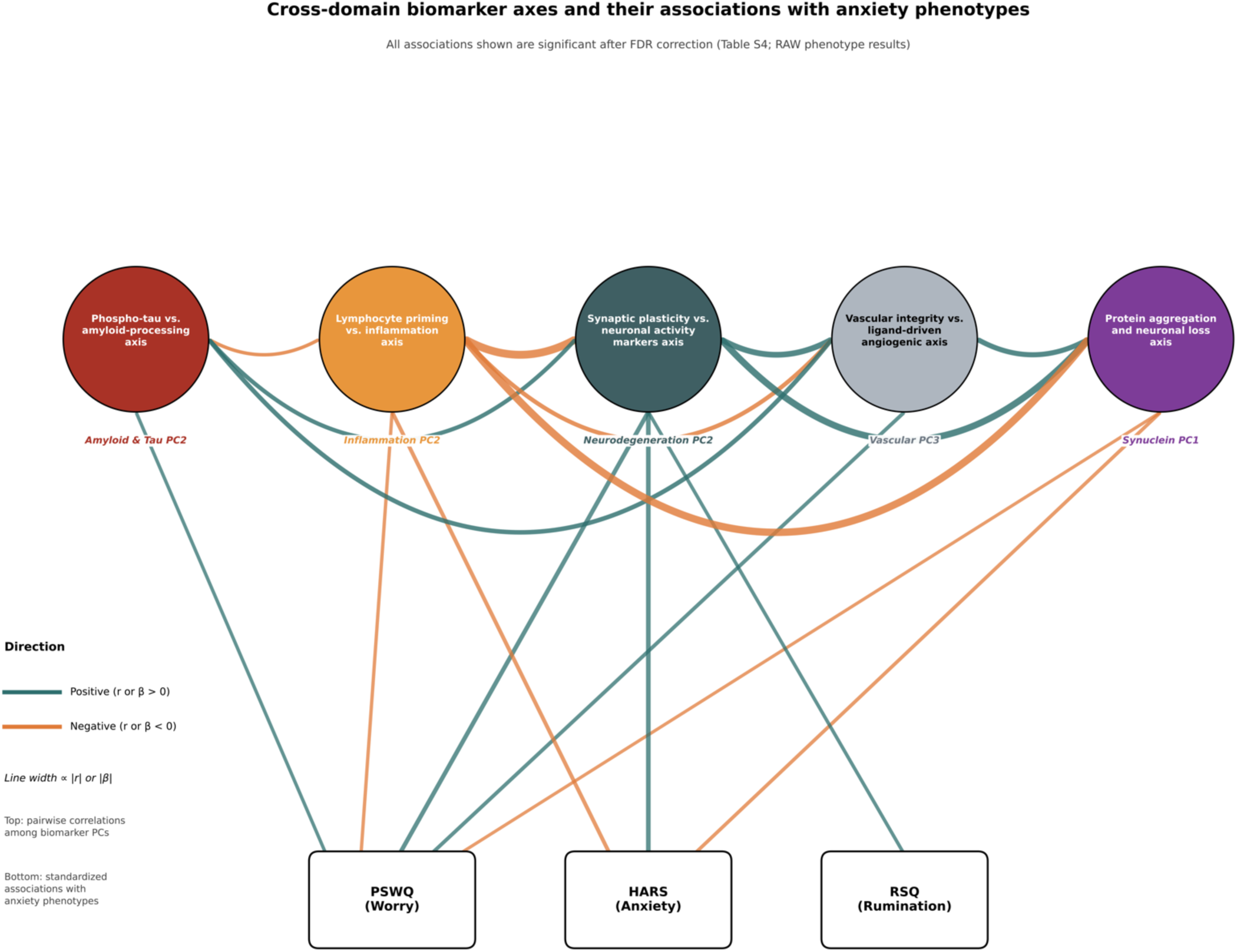
Cross-domain biomarkers axes and their associations with anxiety phenotypes. The circular network displays significant correlations among principal components (PCs). Only correlations that remained significant after false discovery rate (FDR) correction (p-FDR < 0.05) are shown. Edge width and opacity are proportional to the absolute correlation coefficient (|r|), with thicker and darker edges representing stronger correlations. Significance levels are indicated by asterisks next to the node labels.

### A neuronal pentraxin versus neurogranin axis is the most consistent correlate of anxiety phenotypes

The most consistent finding was a neurodegeneration-domain axis on which neuronal pentraxins (NPTX1, NPTX2, NPTXR), neurofilament light (NEFL), and visinin-like protein 1 (VSNL1) loaded positively and neurogranin, annexin A5, FGF2, and enolase 2 loaded negatively (**Supplementary Table S2**). Higher scores (the direction associated with greater worry, anxiety, and rumination) therefore reflect higher pentraxins, NEFL, VSNL1 and lower neurogranin. Several of these proteins are frequently seen as a reactive or compensatory response of synaptic and neuronal injury^30^. Plasma neurofilament light is a validated marker of axonal neurodegeneration^31,32^. Plasma NPTX1, which is induced by low neuronal activity and is proapoptotic, is elevated in mild cognitive impairment and early AD, though the amyloid dependence of this signal has been established mainly in transgenic mouse models rather than in human plasma^33,34^, and NPTX1 contributes to amyloid-β-evoked synaptic damage in cell-based models^34^. VSNL1 rises with neuronal injury and predicts cognitive decline, although that evidence derives from cerebrospinal fluid (CSF)^35^. Their joint elevation at the high-symptom pole is consistent with early synaptic and neuronal injury. NPTX1 and NPTX2 are usually regulated in opposite directions. NPTX1 rises with low neuronal activity and has proapoptotic effects^36^. NPTX2 depends on neuronal activity and declines in AD cerebrospinal fluid^37^. Despite this, both proteins loaded together on the same axis and were associated with high anxiety symptoms. Markers with opposite regulation aligning in the same direction suggest a multifactorial process rather than a single shared mechanism. NPTX2 elevation may instead reflect activity-dependent or disease-related upregulation. This fit reports that NPTX2 increases in synucleinopathy^38^, and matches our finding that this axis correlates with the synuclein domain (**Supplementary Figure S2A**).

Although neurogranin’s CSF elevation indexes synaptic degeneration^39^, it does not track its central pool in plasma^40^, so we do not interpret lower plasma neurogranin as reduced postsynaptic signaling. For these reasons we describe the axis by its markers rather than as a unitary synaptic-dysfunction construct. That it tracked three overlapping but distinct phenotypes, suggests a shared neurodegeneration-domain correlate of anxiety rather than a phenotype-specific one, and within the synuclein domain it co-varied with lower aggregation-prone α-synuclein and TDP-43 markers and higher neurotrophic markers, a cross-domain pattern we report descriptively.

Anxiety associates with hypothalamic-pituitary-adrenal (HPA) activity, glucocorticoid signaling, and neuroinflammatory pathways that, in animal models, produce dendritic remodeling and synapse loss across hippocampal, amygdala, and prefrontal circuits^41^. Similar processes in humans could dysregulate the synaptic and axonal proteins captured by this axis. Anxiety also predicts greater risk of cognitive decline and dementia at the population level^1^, a relationship generally attributed to neurodegeneration broadly rather than to amyloid or tau pathology specifically^42,43^. Consistent with that distinction, a recent meta-analysis found no association between anxiety and amyloid-β and tau in cognitively healthy adults ^44^. Our neurodegeneration-domain axis, built from pentraxins, NEFL, VSNL1, and related synaptic and axonal markers rather than amyloid or tau species, is positioned to capture this non-amyloid pathway. Because the constituent proteins index general neuronal and synaptic integrity rather than anxiety-specific processes, we interpret this axis as a shared correlate and make no claim about causal direction or about preclinical neurodegeneration.

### Dimension-specific biological patterning across inflammatory, vascular, and amyloid-tau axes

The lymphocyte priming versus inflammation axis was associated with worry and global anxiety. Higher symptoms corresponded to the low-score pole, with relatively higher VCAM1, CX3CL1, CRH, IFN-γ, and GFAP and lower CD40L, IL-7, and CCL17, a shift toward an endothelial, astroglial, and stress-marker profile and away from lymphocyte priming, not toward adaptive immune activation. One reading is chronic stress-axis engagement, indexed by higher CRH, with glucocorticoid-associated reducing of lymphocyte signaling^45^, alongside an endothelial and astroglial signature in which CX3CL1 reflects neuron-microglia signaling^46^. The presence of GFAP at this pole is notable, since blood GFAP is an early astrocyte-reactivity marker that increases before cognitive impairment^47,48^, though its loading here was modest and GFAP is strongly age-dependent.

The VEGF (vascular endothelial growth factor) receptor-ligand axis was associated with worry severity. Higher worry corresponded to elevated FLT1 and KDR (also known as VEGFR1 and VEGFR2, respectively) with reduced ligands (VEGFA and PGF). Soluble FLT1 (VEGFR1) binds VEGF-A and placental growth factor (PGF) and restricts their access to signaling receptors, providing negative feedback on VEGF signaling^49^, and a circulating excess of soluble VEGFR1 relative to free ligand has been tied to endothelial dysfunction and impaired angiogenesis in vascular and aging-related states^50,51^. Through interactions with its receptors, VEGF regulates critical pathways such as gene expression, blood-brain barrier function and exhibits neurotrophic and neuroprotective proprieties, collectively promoting neuronal survival^52^. This direction is concordant with the one study testing circulating VEGF against anxiety in non-clinical adults, in which peripheral VEGF was inversely associated with anxiety^53^, though the broader VEGF literature in affective disorders is inconsistent. We describe the axis as a worry-specific imbalance in peripheral VEGF signaling, plausibly endothelial or stress-related, requiring replication before any interpretation can be made involving cerebral blood flow or vascular function. Age is a particular concern for the ligand side of this axis, plasma PGF rises with age in healthy populations, and the high worry group was significantly younger. However, we adjusted for age throughout the analyses, thus we can exclude the possibility that lower PGF could reflect age rather worry severity itself.

The phospho-tau versus amyloid-processing axis was associated with worry only. Higher worry corresponded to lower phospho-tau together with higher Aβ40, Aβ38, and amyloid-processing markers, with Aβ42 contributing minimally (**Supplementary Table S3**). The lower phospho-tau direction runs against the early rise of p-tau217 and p-tau231 in amyloid-onset preclinical AD^54^, and the signal derives from plasma amyloid-processing species rather than amyloid PET, where higher plasma Aβ does not map directly onto greater brain amyloid burden. The specificity of this association to worry in our study, rather than to anxiety as a global construct, invites comparison with a growing literature suggesting that anxiety is not a unitary risk factor for Alzheimer’s disease, but rather that individual symptom dimensions carry distinct biological associations^55^. In cognitively unimpaired older adults, cortical amyloid-β burden measured by PET has been associated specifically with self-reported worry, independent of global anxiety, depression, and cognitive status, with no corresponding association with tau^44^. Our finding that higher phospho-tau versus amyloid-processing axis associated with higher worry, but not global anxiety and rumination, is consistent with this phenotype dissociation, even though the two studies interrogate distinct biological processes, one reflects fibrillar amyloid deposition in brain, and whereas our axis is a plasma composite dominated by phospho-tau and shorter-form amyloid-processing peptides, with Aβ42 contributing minimally to its loading structure. We therefore interpret the convergence between the two studies as support for worry as a specific phenotype that correlates in AD-related biology across measurement modalities, rather than as evidence that the two signals arise from a shared molecular mechanism; establishing the latter would require direct comparison of plasma and PET measures within the same cohort.^20^.

### Symptom chronicity modifies the proteomic signature of anxiety

Stratification by symptom onset duration provided additional context for interpreting the primary findings. The synaptic disorganization axis, the most robust signal in the primary analysis, was attenuated in both chronicity subgroups. However, this attenuation should be interpreted cautiously, the weaker subgroup-specific associations may reflect reduced power rather than the absence of a biological relationship. Nevertheless, differences observed across symptom-duration strata suggest that symptom chronicity may contribute to heterogeneity in the proteomic signature of anxiety. Most notably, the association between the tau-versus-amyloid axis and rumination reversed direction in participants with symptom onset fewer than 10 years prior, such that more recent ruminators showed a profile of elevated tau phosphorylation relative to amyloid - the inverse of the pattern in the full cohort. This reversal may reflect dynamic, stage-dependent reorganization of amyloid-tau relationships during the early anxious prodrome, though replication in larger stratified samples is needed. These findings collectively suggest that the biological correlates of repetitive negative thinking are not static and may evolve with symptom chronicity, with implications for the timing of biomarker-based risk stratification.

### Limitations

Our study has several limitations. First, the cross-sectional analysis does not allow us to draw causal inference. The directionality of observed associations (e.g., whether anxiety drives proteomic dysregulation, early ADRD pathology manifests as anxiety, or both arise from a shared upstream process) cannot be established from a single time point (baseline) of the RAW study. These cross-sectional data were collected within a longitudinal study, and we anticipate that incorporating the 2-year follow-up data in future work will allow us to address directionality. Second, the sample was predominantly White, female, and highly educated, limiting generalizability to more diverse populations. Third, the chronicity stratification subgroups were small (n=19 and n=39) and thus, those results should be considered hypothesis-generating rather than confirmatory. Finally, plasma proteomics reflects peripheral biology and may not fully capture central nervous system processes, though the NULISAseq CNS panel was specifically designed to enrich for CNS-relevant targets with established brain-to-blood correlation.

## CONCLUSION

In at-risk older adults, distinct anxiety dimensions mapped onto dissociable plasma proteomic signatures, with a neuronal pentraxin versus neurogranin axis the only correlate shared across rumination, global anxiety, and worry, and inflammatory, vascular, and amyloid-tau axes tracking global anxiety and worry in dimension-specific ways. Multiplex protein profiling offers a way to examine several of these biological processes simultaneously rather than investigating one analyte at a time. These patterns support anxiety as a biologically heterogeneous correlate in the context of ADRD risk and motivate longitudinal, amyloid-stratified replication to test whether any of these biological axes mark individuals on the AD trajectory.

## Data Availability

The datasets used and analyzed in the current study are available from the corresponding authors upon reasonable request for the sole purpose of replicating the results in this study, provided a data use/sharing agreement is established in agreement with IRB stipulations and applicable laws in the Commonwealth of Pennsylvania and the United States.

## Acknowledgments

We would like to thank participants for their participation in the study as well as study staff from the ARGO Neuroscience of Aging Research Group for their work and support.

## Contributions

Study conception and design: APC, HTK, CA; Data acquisition: APC, MFF, MMG, XZ; Analysis and Interpretation: APC, HTK, CA; Drafting: APC, HTK, CA; and Critical Revision: APC, HTK, MFF, MMG, XZ, DLT, MAB, TKK, CA.

