## Supplemental materials for "Proteomic markers linking anxiety phenotypes to Alzheimer’s disease risk: a multi-analyte proteomic analysis"

**Supplementary data**

**1.Supplementary results**

**Plasma proteomic profiling defines biologically distinct principal components associated with anxiety phenotypes**

*Synaptic plasticity vs. neuronal activity markers axis*

Within the Neurodegeneration domain, PC2 captured a biologically meaningful contrast between postsynaptic calcium signaling and presynaptic pentraxin-mediated synaptic organization. The highest negative loadings on PC2 were neurogranin (NRGN; loading=−0.426), annexin A5 (ANXA5; −0.365), fibroblast growth factor 2 (FGF2; −0.359), and enolase 2 (ENO2; −0.340), representing postsynaptic calcium signaling and activity-dependent plasticity. Positive loadings included neuronal pentraxin 2 (NPTX2; +0.231), neurofilament light chain (NEFL; +0.215), visinin-like protein 1 (VSNL1; +0.206), neuronal pentraxin receptor (NPTXR; +0.195), and neuronal pentraxin 1 (NPTX1). Higher scores on this component therefore reflect a shift toward presynaptic pentraxin activity with concurrent reduction in postsynaptic signaling, consistent with early synaptic disorganization.

*Lymphocyte priming vs. inflammation axis*

Inflammation PC2 (lymphocyte priming vs inflammation axis). The largest positive loadings were CD40 ligand (CD40LG, 0.358), IL-7 (0.327), CCL17 (0.309), CXCL1 (0.277), RUVBL2 (0.268), and IL-1β (IL1B, 0.247), followed by TIMP3 (0.235), PRDX6 (0.231), and CCL13 (0.213). The largest negative loadings were VCAM1 (−0.207), CX3CL1 (−0.189), CRH (−0.156), IL-13 (−0.144), IL-9 (−0.123), IFN-γ (IFNG, −0.116), and GFAP (−0.095). Higher scores index higher lymphocyte-associated markers, consistent with a systemic lymphocyte-priming pattern, whereas lower scores index higher adhesion, astroglial, stress-related (CRH), and effector-cytokine markers, consistent with an inflammatory pattern.

*Vascular integrity vs. ligand-driven angiogenic axis*

Vascular PC3 (Vascular integrity versus ligand-driven angiogenic axis). Positive loadings were FLT1 (0.462), HBA1 (0.360), MME/neprilysin (0.293), KDR (0.279), PDGFRB (0.182), and VEGFD (0.144). Negative loadings were VEGFA (−0.397), PGK1 (−0.389), SAA1 (−0.265), POSTN (−0.186), PTN (−0.119), and PGF (−0.103). Higher scores index elevated VEGF receptors (FLT1 and KDR) with reduced VEGF ligands (VEGFA and PGF) and lower acute-phase inflammatory, and matrix-remodeling markers, that is, a receptor-ligand imbalance in VEGF signaling.

*Phospho-tau versus amyloid-processing axis*

Amyloid and Tau Pathologies PC2 (phospho-tau versus amyloid-processing axis). The largest negative loadings were p-tau217 (−0.405), p-tau231 (−0.380), p-tau181 (−0.364), and total MAPT (−0.296). The largest positive loadings were SFRP1 (0.249), APOE (0.247), KLK6 (0.241), Aβ40 (0.240), and Aβ38 (0.226), with smaller contributions from IGFBP7, ACHE, and BACE1. Higher scores index lower phospho-tau and total tau together with higher Aβ40, Aβ38, and amyloid-processing markers. Aβ42 contributed minimally (0.098), so this axis reflects relative phospho-tau reduction alongside higher shorter-form amyloid peptides and processing markers rather than an Aβ42-defined amyloid-positivity signal.

*Protein aggregation and neuronal loss axis*

Synuclein and Synaptic Disorders PC1 (protein aggregation and neuronal loss axis). This component was unipolar, with all retained markers loading in the same direction, and higher scores index greater burden. The largest loadings were phospho-Ser129-α-synuclein (pSNCA-129, 0.338), total α-synuclein (SNCA, 0.336), oligomeric α-synuclein (0.329), MDH1 (0.308), HTT (0.308), TARDBP (0.293), SOD1 (0.292), ARSA (0.288), phospho-TDP-43 (0.258), and BDNF (0.247). Because the component is unipolar, it indexes overall aggregation and neuronal-loss burden rather than a contrast between two states, and it is not specific to α-synuclein, since huntingtin, TDP-43, and SOD1 load comparably. Synuclein and Synaptic Disorders PC3 (ubiquitin-proteasome and dopaminergic axis). UCHL1 (−0.629), β-synuclein (SNCB, −0.459), and NGF (−0.283) loaded negatively, and PARK7/DJ-1 (+0.340) loaded positively, with DDC and other dopaminergic and lysosomal markers contributing. Higher scores index higher PARK7/DJ-1 relative to UCHL1 and β-synuclein.

**Table S1. List of biomarkers included in the NULISAseq™ CNS Disease Panel 120**

| Biomarker | Protein ID | Protein Name | Gene Name |
|---|---|---|---|
| **Amyloid and Tau Pathologies** | | | |
| ACHE | P22303 | Acetylcholinesterase | ACHE |
| APOE | P02649 | Apolipoprotein E | APOE |
| APOE4 | P02649 | Apolipoprotein E | APOE |
| Aβ38 | P05067 | Amyloid-beta precursor protein | APP |
| Aβ40 | P05067 | Amyloid-beta precursor protein | APP |
| Aβ42 | P05067 | Amyloid-beta precursor protein | APP |
| BACE1 | P56817 | Beta-secretase 1 | BACE1 |
| BASP1 | P80723 | Brain abundant membrane attached signal protein 1 | BASP1 |
| CST3 | P01034 | Cystatin-C | CST3 |
| IGFBP7 | Q16270 | Insulin-like growth factor-binding protein 7 | IGFBP7 |
| KLK6 | Q92876 | Kallikrein-6 | KLK6 |
| MAPT | P10636 | Microtubule-associated protein tau | MAPT |
| PSEN1 | P49768 | Presenilin 1 | PSEN1 |
| p-tau181 | P10636 | Microtubule-associated protein tau | MAPT |
| p-tau217 | P10636 | Microtubule-associated protein tau | MAPT |
| p-tau231 | P10636 | Microtubule-associated protein tau | MAPT |
| SFRP1 | Q8N474 | Secreted frizzled-related protein 1 | SFRP1 |
| **Inflammation** | | | |
| CCL11 | P51671 | Eotaxin | CCL11 |
| CCL13 | Q99616 | C-C motif chemokine 13 | CCL13 |
| CCL17 | Q92583 | C-C motif chemokine 17 | CCL17 |
| CCL2 | P13500 | C-C motif chemokine 2 | CCL2 |
| CCL22 | O00626 | C-C motif chemokine 22 | CCL22 |
| CCL26 | Q9Y258 | C-C motif chemokine 26 | CCL26 |
| CCL3 | P10147 | C-C motif chemokine 3 | CCL3 |
| CCL4 | P13236 | C-C motif chemokine ligand 4 | CCL4 |
| CD40LG | P29965 | CD40 ligand | CD40LG |
| CD63 | P08962 | CD63 antigen | CD63 |
| CHI3L1 | P36222 | Chitinase-3-like protein 1 | CHI3L1 |
| CHIT1 | Q13231 | Chitotriosidase-1 | CHIT1 |
| CNTN2 | Q02246 | Contactin-2 | CNTN2 |
| CRH | P06850 | Corticoliberin | CRH |
| CRP | P02741 | C-reactive protein | CRP |
| CSF2 | P04141 | Granulocyte-macrophage colony-stimulating factor | CSF2 |
| CX3CL1 | P78423 | Fractalkine | CX3CL1 |
| CXCL1 | P09341 | Growth-regulated alpha protein | CXCL1 |
| CXCL10 | P02778 | C-X-C motif chemokine 10 | CXCL10 |

| | | | |
|---|---|---|---|
| CXCL8 | P10145 | Interleukin-8, IL8 | CXCL8 |
| FCN2 | Q15485 | Ficolin-2 | FCN2 |
| GDF15 | Q99988 | Growth/differentiation factor 15 | GDF15 |
| GFAP | P14136 | Glial fibrillary acidic protein | GFAP |
| ICAM1 | P05362 | Intercellular adhesion molecule 1 | ICAM1 |
| IFNG | P01579 | Interferon gamma | IFNG |
| IGF1R | P08069 | Insulin-like growth factor 1 receptor | IGF1R |
| IL10 | P22301 | Interleukin-10 | IL10 |
| IL12p70 | P29459\|P29460 | Interleukin-12 subunit beta\|Interleukin-12 subunit alpha | IL12A\|IL12B |
| IL13 | P35225 | Interleukin-13 | IL13 |
| IL15 | P40933 | Interleukin-15 | IL15 |
| IL16 | Q14005 | Pro-interleukin-16 | IL16 |
| IL17A | Q16552 | Interleukin-17A | IL17A |
| IL18 | Q14116 | Interleukin-18 | IL18 |
| IL1B | P01584 | Interleukin-1 beta | IL1B |
| IL2 | P60568 | Interleukin-2 | IL2 |
| IL33 | O95760 | Interleukin 33 | IL33 |
| IL4 | P05112 | Interleukin-4 | IL4 |
| IL5 | P05113 | Interleukin-5 | IL5 |
| IL6 | P05231 | Interleukin-6 | IL6 |
| IL6R | P08887 | Interleukin-6 receptor subunit alpha | IL6R |
| IL7 | P13232 | Interleukin-7 | IL7 |
| IL9 | P15248 | Interleukin-9 | IL9 |
| PRDX6 | P30041 | Peroxiredoxin-6 | PRDX6 |
| RUVBL2 | Q9Y230 | RuvB like AAA ATPase 2 | RUVBL2 |
| S100A12 | P80511 | Protein S100-A12 | S100A12 |
| S100B | P04271 | S100 calcium binding protein B | S100B |
| SFTPD | P35247 | Pulmonary surfactant-associated protein D | SFTPD |
| SLIT2 | O94813 | Slit homolog 2 protein | SLIT2 |
| TAFA5 | Q7Z5A7 | Chemokine-like protein TAFA-5 | TAFA5 |
| TEK | Q02763 | Angiopoietin-1 receptor | TEK |
| TIMP3 | P35625 | Metalloproteinase inhibitor 3 | TIMP3 |
| TNF | P01375 | Tumor necrosis factor | TNF |
| TREM1 | Q9NP99 | Triggering receptor expressed on myeloid cells 1 | TREM1 |
| TREM2 | Q9NZC2 | Triggering receptor expressed on myeloid cells 2 | TREM2 |
| VCAM1 | P19320 | Vascular cell adhesion protein 1 | VCAM1 |
| **Neurodegeneration** | | | |
| ANXA5 | P08758 | Annexin A5 | ANXA5 |
| CALB2 | P22676 | Calretinin | CALB2 |
| ENO2 | P09104 | Gamma-enolase | ENO2 |

| | | | |
|---|---|---|---|
| FGF2 | P09038 | Fibroblast growth factor 2 | FGF2 |
| GDI1 | P31150 | Rab GDP dissociation inhibitor alpha | GDI1 |
| GDNF | P39905 | Glial cell line-derived neurotrophic factor | GDNF |
| GOT1 | P17174 | Aspartate aminotransferase, cytoplasmic | GOT1 |
| MSLN | Q13421 | Mesothelin | MSLN |
| NEFH | P12036 | Neurofilament heavy polypeptide | NEFH |
| NEFL | P07196 | Neurofilament light polypeptide | NEFL |
| NPTX1 | Q15818 | Neuronal pentraxin-1 | NPTX1 |
| NPTX2 | P47972 | Neuronal pentraxin-2 | NPTX2 |
| NPTXR | O95502 | Neuronal pentraxin receptor | NPTXR |
| NPY | P01303 | Neuropeptide Y | NPY |
| NRGN | Q92686 | Neurogranin | NRGN |
| PDLIM5 | Q96HC4 | PDZ and LIM domain 5 | PDLIM5 |
| REST | Q13127 | RE1 silencing transcription factor | REST |
| SMOC1 | Q9H4F8 | SPARC-related modular calcium-binding protein 1 | SMOC1 |
| SNAP25 | P60880 | Synaptosomal-associated protein 25 | SNAP25 |
| SQSTM1 | Q13501 | Sequestosome-1 | SQSTM1 |
| UBB | NA | NA | NA |
| VSNL1 | P62760 | Visinin-like protein 1 | VSNL1 |
| YWHAG | P61981 | Tyrosine 3-monooxygenase/tryptophan 5-monooxygenase activation protein gamma | YWHAG |
| YWHAZ | P63104 | 14-3-3 protein zeta/delta | YWHAZ |
| **Synuclein and Synaptic Disorders** | | | |
| AGRN | O00468 | Agrin | AGRN |
| ARSA | P15289 | Arylsulfatase A | ARSA |
| BDNF | P23560 | Brain-derived neurotrophic factor | BDNF |
| DDC | NA | NA | NA |
| FABP3 | P05413 | Fatty acid-binding protein, heart | FABP3 |
| FOLR1 | P15328 | Folate receptor alpha | FOLR1 |
| HTT | P42858 | Huntingtin | HTT |
| MDH1 | P40925 | Malate dehydrogenase, cytoplasmic | MDH1 |
| NGF | P01138 | Beta-nerve growth factor | NGF |
| Oligo-SNCA | P37840 | Alpha-synuclein | SNCA |
| PARK7 | Q99497 | Protein/nucleic acid deglycase DJ-1 | PARK7 |
| pSNCA-129 | P37840 | Alpha-synuclein | SNCA |
| pTDP43-409 | Q13148 | TAR DNA-binding protein 43 | TARDBP |
| SNCA | P37840 | Alpha-synuclein | SNCA |

| | | | |
|---|---|---|---|
| SNCB | Q16143 | Synuclein beta | SNCB |
| SOD1 | P00441 | Superoxide dismutase [Cu-Zn] | SOD1 |
| TARDBP | Q13148 | TAR DNA-binding protein 43 | TARDBP |
| UCHL1 | P09936 | Ubiquitin carboxyl-terminal hydrolase isozyme L1 | UCHL1 |
| VGF | O15240 | VGF nerve growth factor inducible | VGF |
| **Vascular** | | | |
| HBA1 | P69905 | Hemoglobin subunit alpha | HBA1 |
| PGF | P49763 | Placenta growth factor | PGF |
| SAA1 | P0DJI8 | Serum amyloid A-1 protein | SAA1 |
| VEGFA | P15692 | Vascular endothelial growth factor A | VEGFA |
| VEGFD | O43915 | Vascular endothelial growth factor D | VEGFD |
| FLT1 | P17948 | Vascular endothelial growth factor receptor 1, VEGFR-1 | FLT1 |
| KDR | P35968 | Vascular endothelial growth factor receptor 2, VEGFR-2 | KDR |
| MME | P08473 | Membrane metalloendopeptidase | MME |
| PDGFRB | P09619 | Platelet-derived growth factor receptor beta | PDGFRB |
| PGK1 | P00558 | Phosphoglycerate kinase 1 | PGK1 |
| POSTN | Q15063 | Periostin | POSTN |
| PTN | P21246 | Pleiotrophin | PTN |

**Table S2. Principal component analysis structure by domain**

| Domain | Analytes (N) | PCs Retained | Cumulative Variance (%) | Variance by PC (%) | Top Loading Variables |
|---|---|---|---|---|---|
| Amyloid and Tau Pathologies | 17 | 2 | 52.4 | PC1: 38.6 PC2: 13.8 | PC1: MAPT, Aβ40, Aβ42, IGFBP7, p-tau231, Aβ38, CST3, KLK6, p-tau181, p-tau217 (neg)<br><br>PC2: p-tau217, p-tau231, p-tau181, MAPT (neg); APOE, KLK6, Aβ40, Aβ38, IGFBP7 (pos) |
| Inflammation | 55 | 3 | 37.4 | PC1: 24.0 PC2: 8.2 PC3: 5.2 | PC1: IL-33, ICAM1, IL-15, IL-16, TAFA5, IL-2, VCAM1, CXCL10, TREM1, CXCL8 (neg)<br><br>PC2: CD40LG, IL-7, CCL17, CXCL1, RUVBL2, IL1B, TIMP3, PRDX6, CCL13 (pos); VCAM1 (neg)<br><br>PC3: IL-6, CRP, CHI3L1, CCL4, CCL3 (pos); CRH, SFTPD, IL4, IL12p70, S100B (neg) |
| Neurodegeneration | 24 | 3 | 47.7 | PC1: 23.1 PC2: 15.7 PC3: 8.8 | PC1: GDI1, REST, NPTX1, YWHAG, SQSTM1, CALB2, NPTXR, NEFL, NPTX2, GOT1 (neg)<br><br>PC2: NRGN, ANXA5, FGF2, ENO2 (neg); NPTX2, NEFL, VSNL1, NPTXR, NPTX1, GOT1 (pos)<br><br>PC3: YWHAZ, GDI1, YWHAG, CALB2, PDLIM5 (neg); NEFH, SQSTM1, NEFL, NPY, MSLN (pos) |
| Synuclein and Synaptic Disorders | 19 | 3 | 64.9 | PC1: 42.6 PC2: 13.4 PC3: 8.9 | PC1: pSNCA-129, SNCA, Oligo-SNCA, MDH1, HTT, TARDBP, SOD, ARSA, pTDP43, BDNF (neg)<br><br>PC2: AGRN, FOLR1, FABP3, NGF, VGF DDC, PARK7 (neg); UCHL1, Oligo-SNCA, TARDBP (pos) |

| | | | | | |
|---|---|---|---|---|---|
| | | | | | PC3: UCHL1, SNCB, NGF, pTDP43, DDC, FOLR1 (neg); PARK7, ARSA, BDNF, SOD1 (pos) |
| Vascular | 12 | 3 | 48.3 | PC1: 21.9<br>PC2: 14.1<br>PC3: 12.3 | PC1: VEGFA, VEGFD, PGF, KDR, PGK1, SAA1, PDGFRB, MME, HBA1 (neg); PTN (pos)<br><br>PC2: POSTN, FLT1, PTN, KDR, PGF, MME, SAA1 (neg); VEGFD, HBA1, PGK1 (pos)<br><br>PC3: FLT1, HBA1, KDR, MME, KDR, PDGFRB, VEGFD (pos); VEGFA, PGK1, SAA1, POSTN (neg) |

Number of retained components determined by Horn's parallel analysis. Variance (%) reported as variance explained by each retained component. Top loading variables listed in order of loading magnitude; negative loadings indicate that higher plasma concentrations correspond to lower PC scores. PCA = principal component analysis.

**Table S3. Loading of plasma biomarkers across principal components by domain**

| Domain | Biomarker | PC1 | PC2 | PC3 | Average Rank |
|---|---|---|---|---|---|
| **Amyloid and Tau Pathologies** | MAPT | 0.314604 | 0.296253 | | 3 |
| | p-tau231 | 0.291647 | 0.380035 | | 4 |
| | Aβ40 | 0.309412 | 0.240307 | | 5 |
| | p-tau181 | 0.251579 | 0.405136 | | 6 |
| | p-tau217 | 0.279185 | 0.364076 | | 6 |
| | IGFBP7 | 0.292640 | 0.197629 | | 7 |
| | Aβ38 | 0.291628 | 0.226173 | | 8 |
| | KLK6 | 0.279235 | 0.241410 | | 8 |
| | SFRP1 | 0.122261 | 0.248910 | | 10 |
| | Aβ42 | 0.301186 | 0.098445 | | 10 |
| | APOE | 0.012388 | 0.246649 | | 11 |
| | CST3 | 0.289487 | 0.128097 | | 12 |
| | ACHE | 0.207557 | 0.176043 | | 12 |
| | BACE1 | 0.216632 | 0.163155 | | 12 |
| | PSEN1 | 0.224177 | 0.146018 | | 12 |
| | BASP1 | 0.021329 | 0.129836 | | 15 |
| | APOE4 | 0.000623 | 0.142135 | | 16 |
| **Inflammation** | IL16 | 0.185828 | 0.085961 | 0.153423 | 14 |
| | CX3CL1 | 0.162722 | 0.189384 | 0.091923 | 16 |
| | CCL13 | 0.154592 | 0.212893 | 0.102384 | 16 |
| | IL12p70 | 0.149508 | 0.088573 | 0.201018 | 18 |
| | VCAM1 | 0.179816 | 0.206992 | 0.068503 | 18 |
| | CCL17 | 0.123012 | 0.309303 | 0.093664 | 19 |
| | IGF1R | 0.165304 | 0.106774 | 0.076681 | 20 |
| | CCL3 | 0.130223 | 0.088871 | 0.199356 | 20 |
| | TAFA5 | 0.184609 | 0.057678 | 0.089040 | 21 |
| | IL7 | 0.133096 | 0.326592 | 0.068804 | 22 |
| | TREM1 | 0.172776 | 0.041865 | 0.105728 | 22 |
| | CRH | 0.042904 | 0.155821 | 0.303680 | 22 |
| | CNTN2 | 0.118388 | 0.092190 | 0.156535 | 23 |
| | IL15 | 0.187359 | 0.104360 | 0.024154 | 23 |
| | IL1B | 0.146622 | 0.247368 | 0.047002 | 24 |
| | IL13 | 0.093069 | 0.144173 | 0.135983 | 24 |
| | IL33 | 0.201929 | 0.059581 | 0.063296 | 24 |
| | CCL2 | 0.153738 | 0.031146 | 0.184980 | 24 |
| | TIMP3 | 0.113250 | 0.234729 | 0.081112 | 24 |
| | IL18 | 0.160288 | 0.038870 | 0.117649 | 25 |

| | | | | | |
|---|---|---|---|---|---|
| | CD40LG | 0.101027 | 0.358051 | 0.074646 | 25 |
| | SLIT2 | 0.160220 | 0.050708 | 0.102147 | 25 |
| | PRDX6 | 0.093773 | 0.230907 | 0.091213 | 25 |
| | TNF | 0.163047 | 0.060885 | 0.071782 | 26 |
| | CCL11 | 0.160522 | 0.020268 | 0.120570 | 26 |
| | CXCL10 | 0.178012 | 0.065637 | 0.060030 | 26 |
| | CHI3L1 | 0.082226 | 0.074383 | 0.276121 | 26 |
| | IL4 | 0.124695 | 0.021427 | 0.217265 | 26 |
| | CCL26 | 0.092492 | 0.094337 | 0.131459 | 27 |
| | CXCL8 | 0.165951 | 0.079690 | 0.027200 | 28 |
| | RUVBL2 | 0.060684 | 0.268190 | 0.079895 | 28 |
| | SFTPD | 0.134509 | 0.010750 | 0.230904 | 28 |
| | S100B | 0.121756 | 0.020545 | 0.200638 | 29 |
| | TEK | 0.160942 | 0.039295 | 0.071233 | 29 |
| | IL6R | 0.139451 | 0.109523 | 0.025837 | 30 |
| | IL9 | 0.115822 | 0.122706 | 0.051543 | 31 |
| | ICAM1 | 0.191937 | 0.003050 | 0.070549 | 31 |
| | GFAP | 0.141980 | 0.095170 | 0.023504 | 31 |
| | IL6 | 0.111681 | 0.007400 | 0.349811 | 31 |
| | CRP | 0.034984 | 0.035774 | 0.330725 | 32 |
| | CHIT1 | 0.147540 | 0.072094 | 0.029915 | 32 |
| | S100A12 | 0.115835 | 0.015390 | 0.160281 | 32 |
| | CSF2 | 0.124512 | 0.055596 | 0.074628 | 33 |
| | TREM2 | 0.124597 | 0.014408 | 0.106042 | 33 |
| | IL2 | 0.182348 | 0.030584 | 0.018785 | 33 |
| | IL10 | 0.104823 | 0.040096 | 0.083110 | 35 |
| | IFNG | 0.109509 | 0.115884 | 0.010741 | 36 |
| | CCL22 | 0.157774 | 0.018882 | 0.040523 | 36 |
| | CCL4 | 0.059422 | 0.013296 | 0.202886 | 36 |
| | CXCL1 | 0.052059 | 0.277495 | 0.010698 | 36 |
| | IL5 | 0.091445 | 0.101091 | 0.036425 | 36 |
| | CD63 | 0.119917 | 0.013170 | 0.081103 | 38 |
| | GDF15 | 0.063315 | 0.055188 | 0.034603 | 43 |
| | FCN2 | 0.025986 | 0.077968 | 0.004290 | 45 |
| | IL17A | 0.033435 | 0.016345 | 0.002635 | 52 |
| **Neurodegeneration** | YWHAG | 0.262706 | 0.159508 | 0.361932 | 7 |
| | NEFL | 0.248705 | 0.214651 | 0.233075 | 7 |
| | GDI1 | 0.281174 | 0.077453 | 0.363807 | 7 |
| | SQSTM1 | 0.260146 | 0.156333 | 0.285103 | 8 |
| | NPTX1 | 0.265021 | 0.186868 | 0.127213 | 9 |

| | | | | |
|---|---|---|---|---|
| | ANXA5 | 0.194231 | 0.364610 | 0.118040 | 10 |
| | NRGN | 0.149320 | 0.425787 | 0.150212 | 10 |
| | MSLN | 0.206176 | 0.170444 | 0.177304 | 11 |
| | NPTX2 | 0.244045 | 0.230537 | 0.109818 | 11 |
| | CALB2 | 0.252140 | 0.058884 | 0.234683 | 11 |
| | NEFH | 0.208036 | 0.126236 | 0.290514 | 11 |
| | REST | 0.266078 | 0.041451 | 0.137240 | 12 |
| | FGF2 | 0.139350 | 0.358867 | 0.133756 | 13 |
| | NPTXR | 0.251753 | 0.194977 | 0.016871 | 13 |
| | VSNL1 | 0.191153 | 0.205681 | 0.090663 | 14 |
| | YWHAZ | 0.157331 | 0.001866 | 0.423872 | 14 |
| | ENO2 | 0.144257 | 0.340262 | 0.097135 | 14 |
| | GOT1 | 0.236678 | 0.179333 | 0.067501 | 14 |
| | NPY | 0.098081 | 0.135047 | 0.220534 | 15 |
| | PDLIM5 | 0.002186 | 0.165526 | 0.168495 | 15 |
| | SNAP25 | 0.175968 | 0.149725 | 0.113163 | 16 |
| | UBB | 0.166921 | 0.060624 | 0.144915 | 16 |
| | SMOC1 | 0.151109 | 0.129462 | 0.085015 | 19 |
| | GDNF | 0.014748 | 0.005481 | 0.091884 | 22 |
| **Synuclein and Synaptic Disorders** | PARK7 | 0.200319 | 0.185575 | 0.339978 | 7 |
| | UCHL1 | 0.066696 | 0.193241 | 0.629380 | 8 |
| | pTDP43 | 0.258132 | 0.104058 | 0.224540 | 8 |
| | FOLR1 | 0.067838 | 0.464859 | 0.157879 | 8 |
| | NGF | 0.062343 | 0.297116 | 0.283327 | 8 |
| | ARSA | 0.288451 | 0.071980 | 0.176301 | 9 |
| | SNCB | 0.139453 | 0.056247 | 0.459185 | 10 |
| | TARDBP | 0.293296 | 0.102464 | 0.045853 | 10 |
| | AGRN | 0.090671 | 0.529043 | 0.030955 | 10 |
| | HTT | 0.307998 | 0.077984 | 0.040503 | 10 |
| | Oligo.SNCA | 0.329450 | 0.109686 | 0.002743 | 10 |
| | DDC | 0.002528 | 0.189682 | 0.207388 | 11 |
| | pSNCA | 0.338152 | 0.039008 | 0.031777 | 11 |
| | BDNF | 0.246825 | 0.064464 | 0.127000 | 11 |
| | FABP3 | 0.094850 | 0.451642 | 0.016384 | 11 |
| | SNCA | 0.336352 | 0.017513 | 0.042968 | 11 |
| | VGF | 0.048044 | 0.222510 | 0.103599 | 11 |
| | SOD1 | 0.292091 | 0.016292 | 0.113799 | 12 |
| | MDH1 | 0.308185 | 0.021362 | 0.009467 | 13 |
| **Vascular** | VEGFA | 0.449147 | 0.066929 | 0.397416 | 5 |
| | FLT1 | 0.162404 | 0.384952 | 0.462202 | 5 |

| | | | | | |
|---|---|---|---|---|---|
| | PTN | 0.328202 | 0.386601 | 0.119273 | 5 |
| | VEGFD | 0.399727 | 0.376077 | 0.144071 | 5 |
| | KDR | 0.310901 | 0.243319 | 0.278749 | 6 |
| | PGK1 | 0.290370 | 0.237967 | 0.389011 | 6 |
| | POSTN | 0.174413 | 0.467940 | 0.186446 | 6 |
| | HBA1 | 0.171831 | 0.307194 | 0.359997 | 7 |
| | MME | 0.190531 | 0.188590 | 0.293136 | 8 |
| | PGF | 0.304407 | 0.265828 | 0.103270 | 8 |
| | SAA1 | 0.257523 | 0.143944 | 0.265198 | 8 |
| | PDGFRB | 0.261132 | 0.017874 | 0.182028 | 9 |

**Table S4. Significant pairwise Pearson correlations among domain-specific biomarker principal components (FDR q < 0.05)**

| PC 1 | PC 2 | r | p | q (FDR) |
|---|---|---|---|---|
| Amyloid_and_Tau_Pathologies_PC1 | Neurodegeneration_PC1 | 0.848 | < 0.001 | p_FDR < 0.001 |
| Inflammation_PC1 | Neurodegeneration_PC1 | 0.829 | < 0.001 | p_FDR < 0.001 |
| Amyloid_and_Tau_Pathologies_PC1 | Inflammation_PC1 | 0.826 | < 0.001 | p_FDR < 0.001 |
| Inflammation_PC1 | Vascular_PC1 | 0.825 | < 0.001 | p_FDR < 0.001 |
| Inflammation_PC2 | Neurodegeneration_PC2 | -0.740 | < 0.001 | p_FDR < 0.001 |
| Inflammation_PC1 | Synuclein_and_Synaptic_Disorders_PC2 | 0.731 | < 0.001 | p_FDR < 0.001 |
| Amyloid_and_Tau_Pathologies_PC1 | Synuclein_and_Synaptic_Disorders_PC2 | 0.727 | < 0.001 | p_FDR < 0.001 |
| Inflammation_PC2 | Synuclein_and_Synaptic_Disorders_PC1 | -0.685 | < 0.001 | p_FDR < 0.001 |
| Neurodegeneration_PC1 | Vascular_PC1 | 0.683 | < 0.001 | p_FDR < 0.001 |
| Synuclein_and_Synaptic_Disorders_PC2 | Vascular_PC1 | 0.660 | < 0.001 | p_FDR < 0.001 |
| Amyloid_and_Tau_Pathologies_PC1 | Vascular_PC1 | 0.648 | < 0.001 | p_FDR < 0.001 |
| Neurodegeneration_PC2 | Synuclein_and_Synaptic_Disorders_PC1 | 0.596 | < 0.001 | p_FDR < 0.001 |
| Neurodegeneration_PC1 | Synuclein_and_Synaptic_Disorders_PC2 | 0.588 | < 0.001 | p_FDR < 0.001 |
| Neurodegeneration_PC1 | Synuclein_and_Synaptic_Disorders_PC1 | 0.564 | < 0.001 | p_FDR < 0.001 |
| Neurodegeneration_PC2 | Synuclein_and_Synaptic_Disorders_PC2 | -0.524 | < 0.001 | p_FDR < 0.001 |
| Neurodegeneration_PC2 | Vascular_PC3 | 0.510 | < 0.001 | p_FDR < 0.001 |
| Inflammation_PC1 | Synuclein_and_Synaptic_Disorders_PC1 | 0.501 | < 0.001 | p_FDR < 0.001 |
| Synuclein_and_Synaptic_Disorders_PC3 | Vascular_PC2 | 0.494 | < 0.001 | p_FDR < 0.001 |
| Synuclein_and_Synaptic_Disorders_PC1 | Vascular_PC3 | 0.440 | < 0.001 | p_FDR < 0.001 |
| Inflammation_PC2 | Synuclein_and_Synaptic_Disorders_PC2 | 0.438 | < 0.001 | p_FDR < 0.001 |
| Synuclein_and_Synaptic_Disorders_PC1 | Vascular_PC1 | 0.429 | < 0.001 | p_FDR < 0.001 |
| Inflammation_PC3 | Neurodegeneration_PC3 | 0.420 | < 0.001 | p_FDR < 0.001 |
| Amyloid_and_Tau_Pathologies_PC1 | Synuclein_and_Synaptic_Disorders_PC1 | 0.399 | < 0.001 | p_FDR < 0.001 |
| Amyloid_and_Tau_Pathologies_PC2 | Vascular_PC1 | -0.380 | < 0.001 | p_FDR < 0.001 |
| Amyloid_and_Tau_Pathologies_PC2 | Vascular_PC3 | 0.373 | < 0.001 | p_FDR < 0.001 |
| Inflammation_PC2 | Vascular_PC3 | -0.348 | < 0.001 | p_FDR < 0.001 |
| Amyloid_and_Tau_Pathologies_PC2 | Neurodegeneration_PC2 | 0.336 | < 0.001 | p_FDR < 0.01 |
| Amyloid_and_Tau_Pathologies_PC2 | Synuclein_and_Synaptic_Disorders_PC3 | 0.329 | < 0.001 | p_FDR < 0.01 |
| Neurodegeneration_PC1 | Vascular_PC3 | 0.320 | < 0.001 | p_FDR < 0.01 |
| Amyloid_and_Tau_Pathologies_PC2 | Synuclein_and_Synaptic_Disorders_PC2 | -0.287 | 0.00238 | p_FDR < 0.01 |
| Amyloid_and_Tau_Pathologies_PC1 | Vascular_PC2 | 0.283 | 0.00279 | p_FDR < 0.01 |
| Amyloid_and_Tau_Pathologies_PC1 | Vascular_PC3 | 0.279 | 0.00313 | p_FDR < 0.01 |
| Amyloid_and_Tau_Pathologies_PC1 | Synuclein_and_Synaptic_Disorders_PC3 | 0.268 | 0.00459 | p_FDR < 0.05 |
| Inflammation_PC2 | Vascular_PC2 | 0.263 | 0.00542 | p_FDR < 0.05 |
| Inflammation_PC3 | Synuclein_and_Synaptic_Disorders_PC3 | 0.261 | 0.00580 | p_FDR < 0.05 |
| Amyloid_and_Tau_Pathologies_PC2 | Inflammation_PC1 | -0.258 | 0.00653 | p_FDR < 0.05 |
| Neurodegeneration_PC1 | Synuclein_and_Synaptic_Disorders_PC3 | 0.256 | 0.00701 | p_FDR < 0.05 |
| Neurodegeneration_PC2 | Vascular_PC1 | -0.244 | 0.01030 | p_FDR < 0.05 |
| Neurodegeneration_PC3 | Vascular_PC2 | -0.219 | 0.02136 | p_FDR < 0.05 |

FDR correction (Benjamini-Hochberg) applied across all pairwise correlations. Intra-domain PC pairs (orthogonal by construction, $r \approx 0$) are excluded. Of comparisons, 39 survived FDR correction. FDR = false discovery rate; PC = principal component.

**Table S5. Worry-associated principal components (PCs) and their cross-domain.**

**PSWQ (Worry)** — PC associations

Std. β · z-scored variables · Adjusted for age + sex · BH-FDR

| | | Effect | | | Inference | | FDR | |
|---|---|---|---|---|---|---|---|---|
| | N | Std. β[1] | SE | 95% CI | p | Sig. | p (FDR)[2] | Sig. (FDR) |
| Neurodegeneration (PC2) | **110** | **0.356** | **0.082** | **[0.195, 0.518]** | **0.0000** | *** | **0.0007** | *** |
| Vascular (PC3) | **110** | **0.293** | **0.085** | **[0.125, 0.460]** | **0.0009** | *** | **0.0120** | * |
| Synuclein and Synaptic Disorders (PC3) | **110** | **0.264** | **0.087** | **[0.093, 0.436]** | **0.0031** | ** | **0.0236** | * |
| Inflammation (PC2) | **110** | **−0.253** | **0.086** | **[-0.421, -0.084]** | **0.0041** | ** | **0.0236** | * |
| Amyloid and Tau Pathologies (PC2) | **110** | **0.250** | **0.086** | **[0.081, 0.418]** | **0.0045** | ** | **0.0236** | * |
| Synuclein and Synaptic Disorders (PC1) | **110** | **0.227** | **0.087** | **[0.056, 0.397]** | **0.0104** | * | **0.0438** | * |
| Vascular (PC1) | 110 | **−0.146** | 0.089 | [-0.321, 0.029] | 0.1051 | | 0.2792 | |
| Synuclein and Synaptic Disorders (PC2) | 110 | **−0.146** | 0.093 | [-0.328, 0.036] | 0.1187 | | 0.2933 | |
| Neurodegeneration (PC3) | 110 | **−0.125** | 0.094 | [-0.309, 0.059] | 0.1867 | | 0.3430 | |
| Neurodegeneration (PC1) | 110 | **0.105** | 0.094 | [-0.078, 0.288] | 0.2648 | | 0.4101 | |
| Vascular (PC2) | 110 | **0.079** | 0.091 | [-0.099, 0.256] | 0.3889 | | 0.5611 | |
| Inflammation (PC3) | 110 | **−0.030** | 0.090 | [-0.207, 0.147] | 0.7423 | | 0.8205 | |
| Inflammation (PC1) | 110 | **−0.027** | 0.095 | [-0.214, 0.160] | 0.7746 | | 0.8341 | |
| Amyloid and Tau Pathologies (PC1) | 110 | **0.023** | 0.095 | [-0.163, 0.209] | 0.8077 | | 0.8345 | |

[1] All continuous variables z-scored prior to modelling.

[2] BH-FDR correction applied across all PC tests within this outcome.

🟡 Amber + bold = FDR < 0.05 · 🔵 Light blue = nominal p < 0.05 · *** p<.001 · ** p<.01 · * p<.05

**Table S6 Rumination-associated principal components (PCs) and their cross-domain.**

### **RSQ (Rumination)** — PC associations

Std. β · z-scored variables · Adjusted for age + sex · BH-FDR

| | | Effect | | | Inference | | FDR | |
|---|---|---|---|---|---|---|---|---|
| | N | Std. β[1] | SE | 95% CI | p | Sig. | p (FDR)[2] | Sig. (FDR) |
| **Neurodegeneration (PC2)** | **110** | **0.257** | **0.090** | **[0.080, 0.433]** | **0.0052** | ** | **0.0243** | * |
| **Inflammation (PC2)** | 110 | **−0.215** | 0.091 | [-0.393, -0.036] | 0.0202 | * | 0.0706 | |
| **Neurodegeneration (PC3)** | 110 | **−0.198** | 0.097 | [-0.389, -0.008] | 0.0437 | * | 0.1312 | |
| **Synuclein and Synaptic Disorders (PC1)** | 110 | **0.151** | 0.092 | [-0.031, 0.332] | 0.1064 | | 0.2792 | |
| **Synuclein and Synaptic Disorders (PC3)** | 110 | **0.136** | 0.094 | [-0.049, 0.320] | 0.1528 | | 0.3430 | |
| **Inflammation (PC3)** | 110 | **−0.127** | 0.094 | [-0.311, 0.056] | 0.1769 | | 0.3430 | |
| **Amyloid and Tau Pathologies (PC2)** | 110 | **0.124** | 0.093 | [-0.058, 0.305] | 0.1840 | | 0.3430 | |
| **Synuclein and Synaptic Disorders (PC2)** | 110 | **−0.127** | 0.097 | [-0.318, 0.063] | 0.1926 | | 0.3430 | |
| **Vascular (PC3)** | 110 | **0.122** | 0.093 | [-0.062, 0.305] | 0.1960 | | 0.3430 | |
| **Vascular (PC1)** | 110 | **−0.109** | 0.094 | [-0.293, 0.075] | 0.2499 | | 0.4101 | |
| **Inflammation (PC1)** | 110 | **−0.080** | 0.099 | [-0.275, 0.115] | 0.4234 | | 0.5736 | |
| **Vascular (PC2)** | 110 | **0.043** | 0.095 | [-0.143, 0.229] | 0.6516 | | 0.7602 | |
| **Neurodegeneration (PC1)** | 110 | **0.023** | 0.098 | [-0.170, 0.216] | 0.8146 | | 0.8345 | |
| **Amyloid and Tau Pathologies (PC1)** | 110 | **−0.018** | 0.099 | [-0.212, 0.177] | 0.8582 | | 0.8582 | |

[1] All continuous variables z-scored prior to modelling.

[2] BH-FDR correction applied across all PC tests within this outcome.

🟡 Amber + bold = FDR < 0.05 · 🔵 Light blue = nominal p < 0.05 · *** p<.001 · ** p<.01 · * p<.05

**Table S7. Global anxiety-associated principal components (PCs) and their cross-domain.**

**HARS (Anxiety)** — PC associations

Std. β · z-scored variables · Adjusted for age + sex · BH-FDR

| | | Effect | | | Inference | | FDR | |
|---|---|---|---|---|---|---|---|---|
| | N | Std. β[1] | SE | 95% CI | p | Sig. | p (FDR)[2] | Sig. (FDR) |
| Neurodegeneration (PC2) | 110 | 0.385 | 0.089 | [0.210, 0.560] | 0.0000 | *** | 0.0007 | *** |
| Inflammation (PC2) | 110 | −0.308 | 0.092 | [-0.489, -0.127] | 0.0011 | ** | 0.0120 | * |
| Synuclein and Synaptic Disorders (PC1) | 110 | 0.273 | 0.093 | [0.090, 0.456] | 0.0042 | ** | 0.0236 | * |
| Synuclein and Synaptic Disorders (PC2) | 110 | −0.235 | 0.099 | [-0.429, -0.041] | 0.0192 | * | 0.0706 | |
| Amyloid and Tau Pathologies (PC2) | 110 | 0.208 | 0.095 | [0.022, 0.393] | 0.0303 | * | 0.0980 | |
| Vascular (PC3) | 110 | 0.138 | 0.097 | [-0.052, 0.327] | 0.1576 | | 0.3430 | |
| Neurodegeneration (PC1) | 110 | 0.113 | 0.101 | [-0.086, 0.312] | 0.2675 | | 0.4101 | |
| Synuclein and Synaptic Disorders (PC3) | 110 | 0.108 | 0.098 | [-0.084, 0.300] | 0.2734 | | 0.4101 | |
| Vascular (PC1) | 110 | −0.082 | 0.098 | [-0.274, 0.109] | 0.4008 | | 0.5611 | |
| Amyloid and Tau Pathologies (PC1) | 110 | −0.078 | 0.103 | [-0.279, 0.123] | 0.4476 | | 0.5875 | |
| Neurodegeneration (PC3) | 110 | 0.067 | 0.102 | [-0.134, 0.268] | 0.5127 | | 0.6334 | |
| Vascular (PC2) | 110 | −0.064 | 0.098 | [-0.256, 0.129] | 0.5202 | | 0.6334 | |
| Inflammation (PC1) | 110 | −0.065 | 0.103 | [-0.267, 0.137] | 0.5278 | | 0.6334 | |
| Inflammation (PC3) | 110 | 0.040 | 0.098 | [-0.152, 0.232] | 0.6832 | | 0.7755 | |

[1] All continuous variables z-scored prior to modelling.

[2] BH-FDR correction applied across all PC tests within this outcome.

🟡 Amber + bold = FDR < 0.05 · 🔵 Light blue = nominal p < 0.05 · *** p<.001 · ** p<.01 · * p<.05

### Table S8. Demographics and Clinical Characteristics of Participants - Onset of Symptoms

**Table 1.** Demographics and Clinical Characteristics of Participants

| Variable | Low-RAW N = 62[1] | High-RAW N = 48[1] | p-value[2] | Overall N = 110[1] |
|---|---|---|---|---|
| ***Age (years)*** | 66.8 ± 8.5 | 61.2 ± 7.6 | **<0.001** | 64.3 ± 8.6 |
| ***Sex*** | | | 0.400 | |
| Male | 24 (39%) | 14 (29%) | | 38 (35%) |
| Female | 38 (61%) | 34 (71%) | | 72 (65%) |
| ***Race*** | | | >0.999 | |
| White | 53 (85%) | 41 (85%) | | 94 (85%) |
| Non-White | 9 (15%) | 7 (15%) | | 16 (15%) |
| ***Ethnicity*** | | | 0.592 | |
| Not Hispanic or Latino | 60 (97%) | 48 (100%) | | 108 (98%) |
| Hispanic or Latino | 2 (3.2%) | 0 (0%) | | 2 (1.8%) |
| ***APOEε4 (carrier)*** | | | 0.568 | |
| Non-carrier | 41 (76%) | 38 (83%) | | 79 (79%) |
| Carrier | 13 (24%) | 8 (17%) | | 21 (21%) |
| ***Education (years)*** | 16.8 ± 2.2 | 16.3 ± 2.2 | 0.329 | 16.6 ± 2.2 |
| ***Onset of symptoms*** | | | 0.099 | |
| Less than 10 years | 6 (60%) | 13 (27%) | | 19 (33%) |
| ≥10 years | 4 (40%) | 35 (73%) | | 39 (67%) |
| ***PSWQ (Worry)*** | 32.2 ± 9.0 | 57.9 ± 11.2 | **<0.001** | 43.4 ± 16.2 |
| ***RSQ (Rumination)*** | 29.6 ± 5.6 | 48.2 ± 11.8 | **<0.001** | 37.7 ± 12.8 |
| ***HARS (Anxiety)*** | 5.5 ± 3.9 | 15.1 ± 7.1 | **<0.001** | 9.7 ± 7.3 |

[1] Mean ± SD; n (%)

[2] Wilcoxon rank sum test; Pearson's Chi-squared test

*Mean ± SD for continuous variables; n (%) for categorical variables.*

*p-values: Wilcoxon rank-sum test (continuous); Pearson Chi-squared test (categorical).*

*Onset of symptoms: collapsed from 4-level scale (1 = <1 yr, 2 = 1–5 yrs, 3 = 5–10 yrs, 4 = ≥10 yrs) into <10 years vs. ≥10 years, using the longest duration reported across PSWQ, RSQ, and HARS.*

**Figure S1. Pairwise Pearson correlations among domain-specific biomarker principal components.** Each cell shows the correlation coefficient between two principal components derived from five biomarker domains: Amyloid and Tau Pathologies, Inflammation, Neurodegeneration, Synuclein and Synaptic Disorders, and Vascular. Within each domain, the first three components (PC1 to PC3) are shown. Cell color encodes the direction and magnitude of the correlation, with orange indicating positive and teal indicating negative associations, scaled from −1.0 to +1.0 (see color bar). Only correlations significant after false discovery rate correction ($p_{FDR} < 0.05$) are displayed; blank cells were not significant at this threshold. Asterisks denote significance levels: *$p_{FDR} < 0.05$, **$p_{FDR} < 0.01$, ***$p_{FDR} < 0.001$.

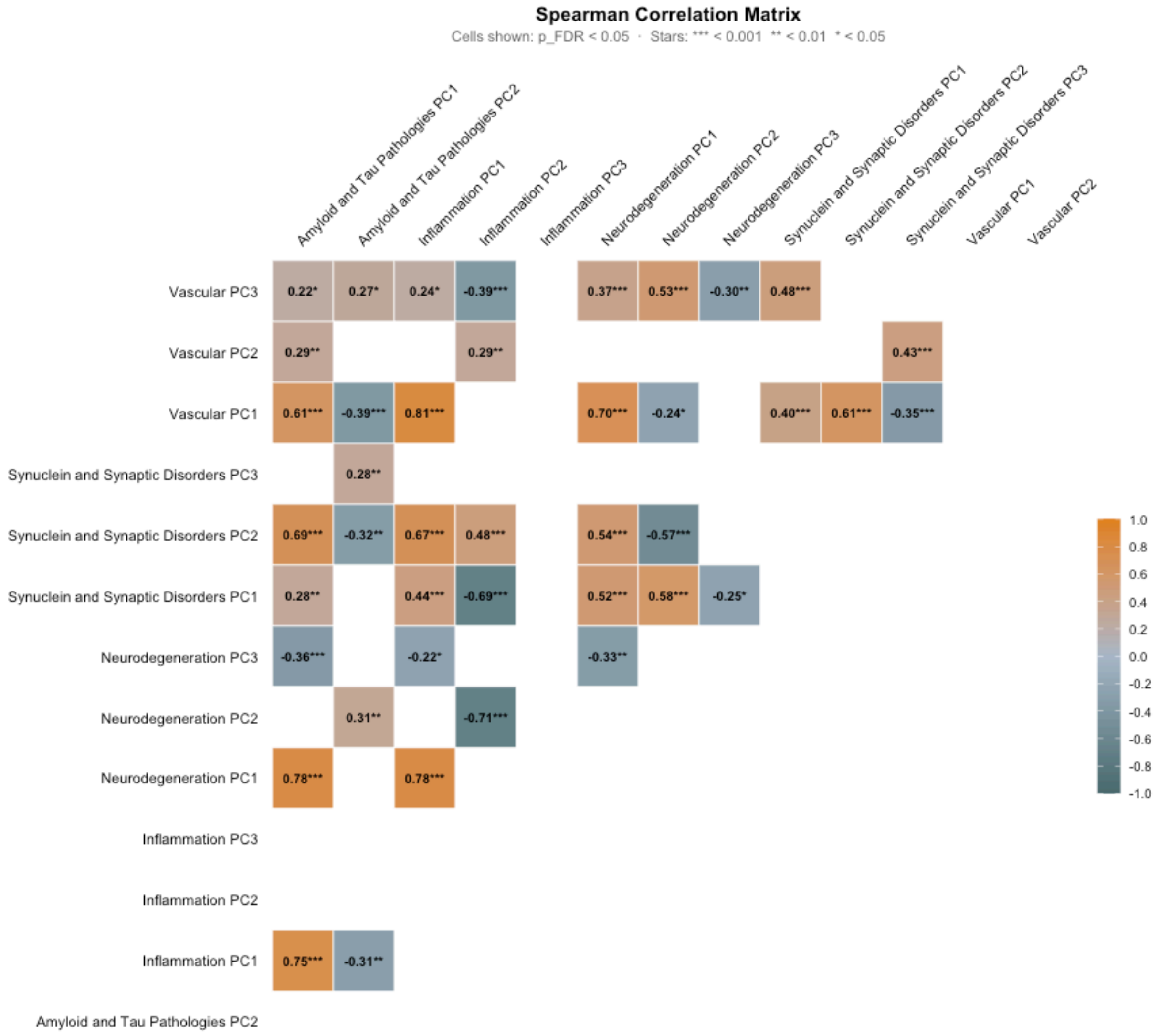
Spearman Correlation Matrix
Cells shown: p_FDR < 0.05 · Stars: *** < 0.001 ** < 0.01 * < 0.05
Amyloid and Tau Pathologies PC1
Amyloid and Tau Pathologies PC2
Inflammation PC1
Inflammation PC2
Inflammation PC3
Neurodegeneration PC1
Neurodegeneration PC2
Neurodegeneration PC3
Synuclein and Synaptic Disorders PC1
Synuclein and Synaptic Disorders PC2
Synuclein and Synaptic Disorders PC3
Vascular PC1
Vascular PC2
Vascular PC3
0.22* 0.27* 0.24* -0.39*** 0.37*** 0.53*** -0.30** 0.48***
Vascular PC2
0.29** 0.29** 0.43***
Vascular PC1
0.61*** -0.39*** 0.81*** 0.70*** -0.24* 0.40*** 0.61*** -0.35***
Synuclein and Synaptic Disorders PC3
0.28**
Synuclein and Synaptic Disorders PC2
0.69*** -0.32** 0.67*** 0.48*** 0.54*** -0.57***
Synuclein and Synaptic Disorders PC1
0.28** 0.44*** -0.69*** 0.52*** 0.58*** -0.25*
Neurodegeneration PC3
-0.36*** -0.22* -0.33**
Neurodegeneration PC2
0.31** -0.71***
Neurodegeneration PC1
0.78*** 0.78***
Inflammation PC3
Inflammation PC2
Inflammation PC1
0.75*** -0.31**
Amyloid and Tau Pathologies PC2
1.0
0.8
0.6
0.4
0.2
0.0
-0.2
-0.4
-0.6
-0.8
-1.0

**Figure S2. Correlation network of plasma proteomics by domain.** Nodes represent the principal components (PCs), colored by domain. Each panel (A-E) corresponds to one biological axis. Edges are pairwise Pearson correlations significant after Benjamini-Hochberg correction (p-FDR≤0.0021). Edge color denotes direction (teal, positive; orange, negative) and edge width/opacity scale with |r|. FDR-adjusted significance (*p<0.05, **p<0.01, ***p<0.001).

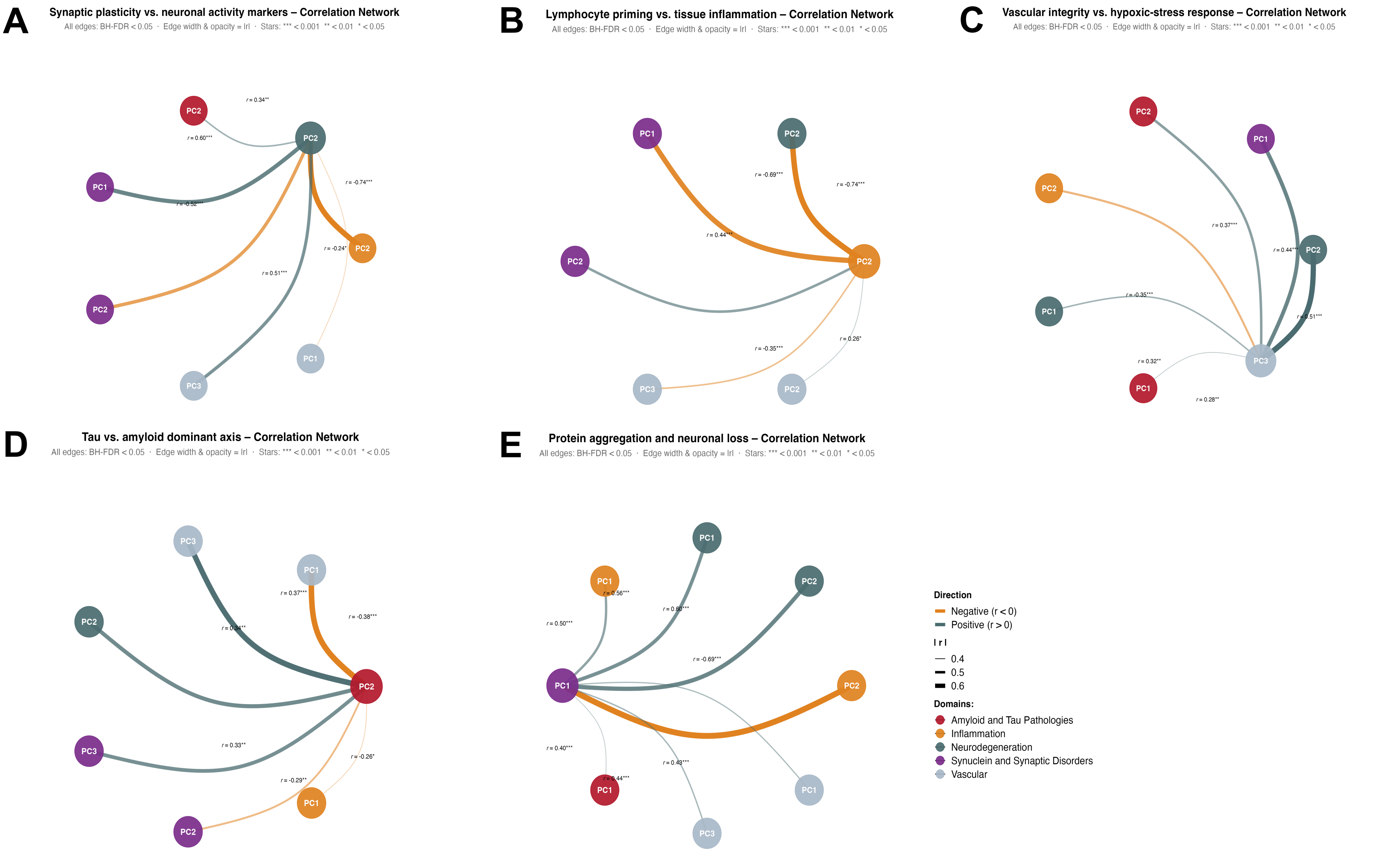

**Figure S3. Volcano plot of principal component (PC) associations with anxiety phenotypes.** Each point represents linear regression tests (14 PCs × 3 anxiety phenotypes), by phenotype: worry (PSWQ), anxiety (HARS), and rumination (RSQ). The x-axis shows the standardized regression coefficient (β; direction and magnitude of association) and the y-axis shows statistical significance as $-\log_{10}$(BH-FDR-adjusted p). The dashed horizontal line marks the FDR=0.05 threshold; points above it (filled, colored, and labeled with the PC number) are statistically significant.

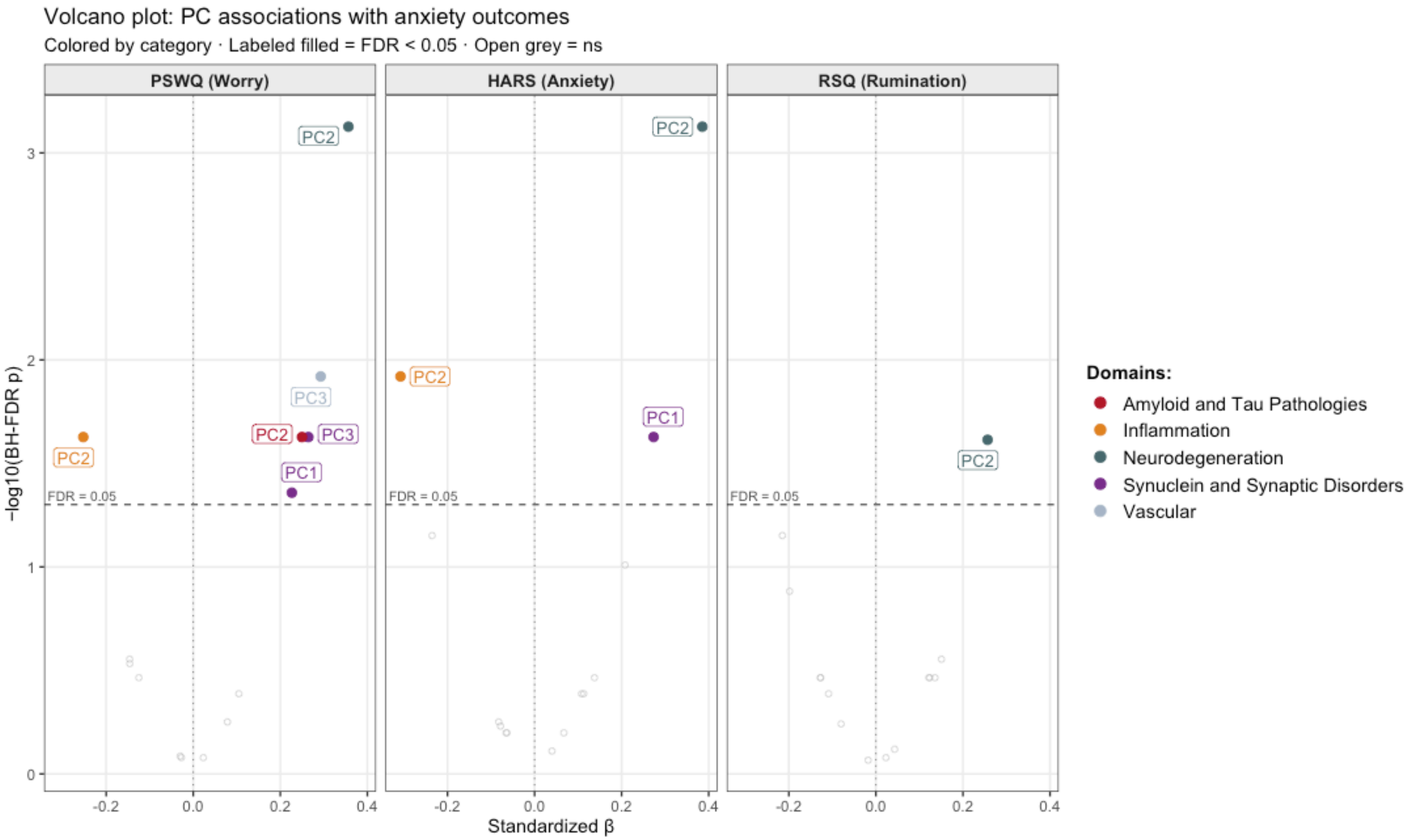

**Figure S4. Anxiety-associated principal components (PCs) and their cross-domain correlates by onset of symptoms.** Panel A corresponds to one biological axis significantly associated with one or more anxiety phenotypes with onset of symptoms $< 10$ years. Panel B corresponds to one biological axis significantly associated with one or more anxiety phenotypes with onset of symptoms $\geq 10$ years. Scatterplots of the PCs against anxiety phenotype, with linear regression fit (solid line) and 95% confidence band (shaded). All models were adjusted for age and sex ($N = 110$); standardized $\beta$, FDR-adjusted p-value, and model $R^2$ are shown.

**A. Onset of symptoms <10 years**

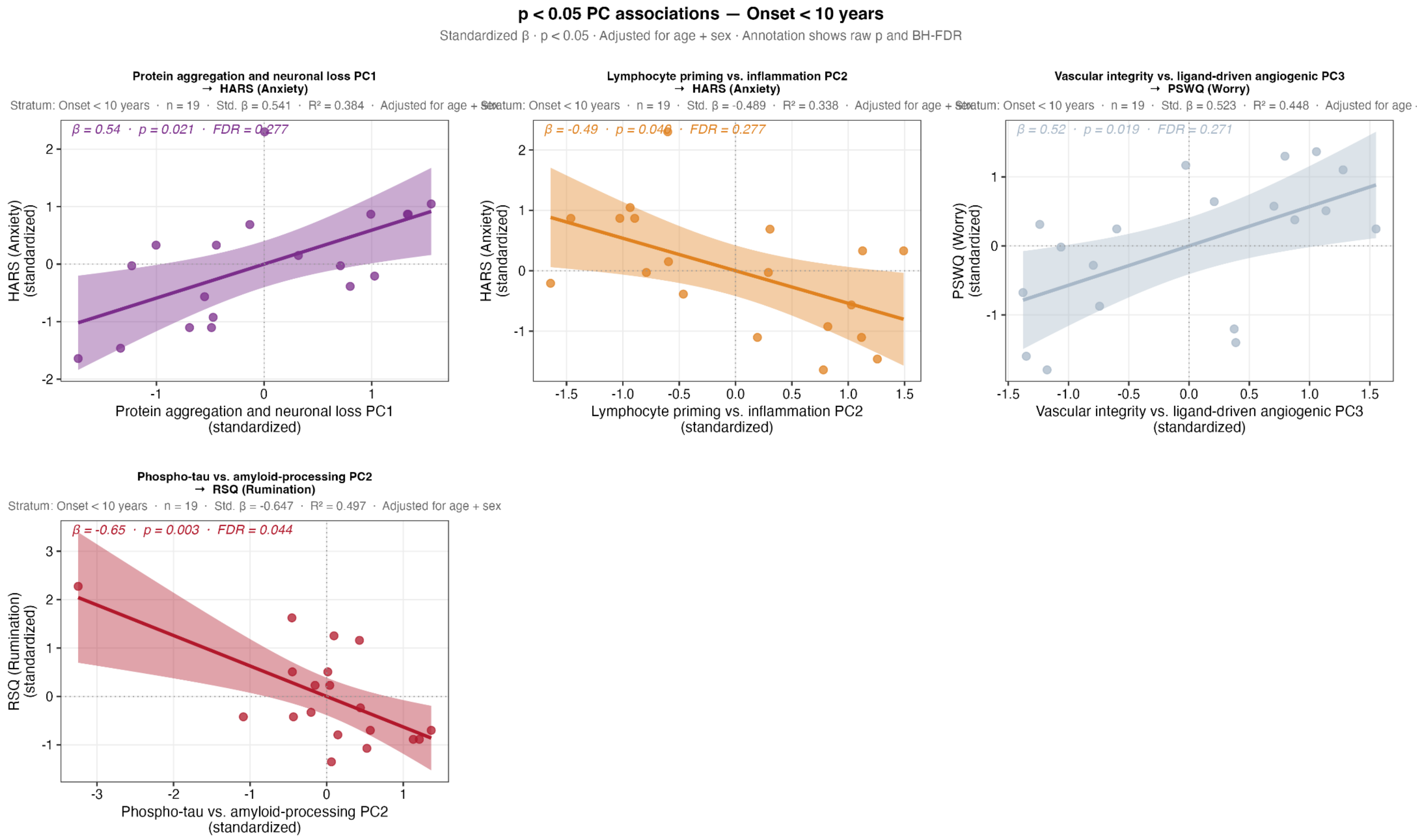
p < 0.05 PC associations — Onset < 10 years
Standardized β · p < 0.05 · Adjusted for age + sex · Annotation shows raw p and BH-FDR
Protein aggregation and neuronal loss PC1
→ HARS (Anxiety)
Stratum: Onset < 10 years · n = 19 · Std. β = 0.541 · R² = 0.384 · Adjusted for age + sex
β = 0.54 · p = 0.021 · FDR = 0.277
HARS (Anxiety)
(standardized)
Protein aggregation and neuronal loss PC1
(standardized)
Lymphocyte priming vs. inflammation PC2
→ HARS (Anxiety)
Stratum: Onset < 10 years · n = 19 · Std. β = -0.489 · R² = 0.338 · Adjusted for age + sex
β = -0.49 · p = 0.046 · FDR = 0.277
HARS (Anxiety)
(standardized)
Lymphocyte priming vs. inflammation PC2
(standardized)
Vascular integrity vs. ligand-driven angiogenic PC3
→ PSWQ (Worry)
Stratum: Onset < 10 years · n = 19 · Std. β = 0.523 · R² = 0.448 · Adjusted for age ·
β = 0.52 · p = 0.019 · FDR = 0.271
PSWQ (Worry)
(standardized)
Vascular integrity vs. ligand-driven angiogenic PC3
(standardized)
Phospho-tau vs. amyloid-processing PC2
→ RSQ (Rumination)
Stratum: Onset < 10 years · n = 19 · Std. β = -0.647 · R² = 0.497 · Adjusted for age + sex
β = -0.65 · p = 0.003 · FDR = 0.044
RSQ (Rumination)
(standardized)
Phospho-tau vs. amyloid-processing PC2
(standardized)

### B. Onset of symptoms ≥10 years

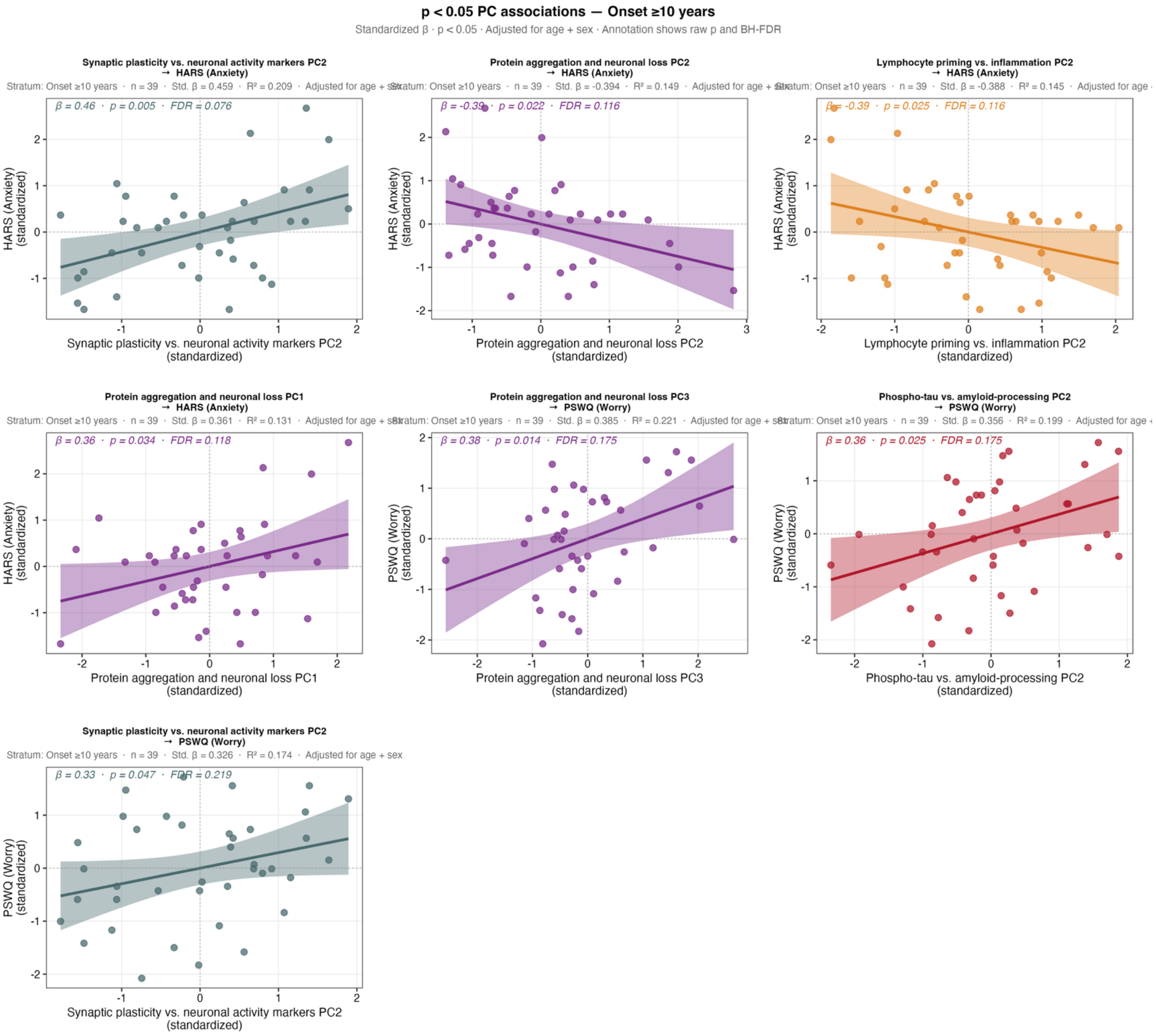
